# Interpreting convolutional neural network explainability for head-and-neck cancer radiotherapy organ-at-risk segmentation

**DOI:** 10.1101/2025.07.30.25332421

**Authors:** Victor I. J. Strijbis, O.J. Gurney-Champion, Dragos I. Grama, Berend J. Slotman, Wilko F.A.R. Verbakel

## Abstract

**Background:** Convolutional neural networks (CNNs) have emerged to reduce clinical resources and standardize auto-contouring of organs-at-risk (OARs). Although CNNs perform adequately for most patients, understanding when the CNN might fail is critical for effective and safe clinical deployment. However, the limitations of CNNs are poorly understood because of their *black-box* nature. Explainable artificial intelligence (XAI) can expose CNNs’ inner mechanisms for *classification*. Here, we investigate the inner mechanisms of CNNs for *segmentation* and explore a novel, computational approach to *a-priori* flag potentially insufficient parotid gland (PG) contours.

**Methods:** First, 3D UNets were trained in three PG segmentation situations using (1) synthetic cases; (2) 1925 clinical computed tomography (CT) scans with typical and (3) more consistent contours curated through a previously validated auto-curation step. Then, we generated attribution maps for seven XAI methods, and qualitatively assessed them for congruency between simulated and clinical contours, and how much XAI agreed with expert reasoning. To objectify observations, we explored persistent homology intensity filtrations to capture essential topological characteristics of XAI attributions. Principal component (PC) eigenvalues of Euler characteristic profiles were correlated with spatial agreement (Dice-Sørensen similarity coefficient; DSC). Evaluation was done using sensitivity, specificity and the area under receiver operating characteristic (AUROC) curve on an external AAPM dataset, where as proof-of-principle, we regard the lowest 15% DSC as insufficient.

**Results:** PatternNet attributions (PNet-A) focused on soft-tissue structures, whereas guided backpropagation (GBP) highlighted both soft-tissue and high-density structures (e.g. mandible bone), which was congruent with synthetic situations. Both methods typically had higher/denser activations in better auto-contoured medial and anterior lobes. Curated models produced “cleaner” gradient class-activation mapping (GCAM) attributions. Quantitative analysis showed that PCλ_1_ of guided GCAM’s (GGCAM) Euler characteristic (EC) profile had good predictive value (sensitivity>0.85, specificity>0.9) of DSC for AAPM cases, with AUROC=0.66, 0.74, 0.94, 0.83 for GBP, GCAM, GGCAM and PNet-A. For for λ_1_<-1.8e3 of GGCAM’s EC-profile, 87% of cases were insufficient.

**Conclusions:** GBP and PNet-A qualitatively agreed most with expert reasoning on directly (structure borders) and indirectly (proxies used for identifying structure borders) important features for PG segmentation. Additionally, this work investigated as proof-of-principle how topological data analysis could possibly be used for quantitative XAI signal analysis to *a-priori* mark potentially inadequate CNN-segmentations, using only features from inside the predicted PG. This work used PG as a well-understood segmentation paradigm and may extend to target volumes and other organs-at-risk.

## 1. Introduction

In radiation oncology, the radiotherapy target and organ-at-risk structures have a crucial role in that they determine where ionizing radiation is directed, and which areas are spared. Manual segmentation (or contouring) of these structures, however, is a highly time-consuming procedure that introduces inter- and intra-rater variabilities due to low image contrast, scattering noise, imaging artefacts, rater experience, anatomical limitations, bias and human errors. Convolutional neural networks (CNNs) have emerged to reduce resource allocation and improve consistency for segmentation.

The increasing complexity of CNNs accompanies a *black-box-*nature model that lacks transparency and interpretability and thereby complicates understanding when the CNN might give incorrect results, which holds back the adoption of deep learning in the medical domain^1–3^. In addition, like human segmentation, CNN-based segmentation can deliver less accurate segmentations and identifying such cases is difficult because the limitations of CNNs are not well understood. Hence, for safe and reliable deployment of CNNs in clinic, it is desirable to have explainable artificial intelligence (XAI) methods that explain how segmentations were achieved. This may aid in the detection of sub-optimal CNN contours and build trust in CNNs by exposing causal relationships between input features and CNN output^4–7^.

XAI includes *global* and *local* explanation methods. *Global* explanations visualize the (C)NN’s filters learned from the *global* training dataset, whereas *local* explanations highlight features (both filters and individual voxel features) that were important in the CNN’s decision-making, specifically for individual cases. Since, for radiation oncology, the goal of XAI is to expose important features for a CNN’s decision in individual patients (saliency mapping), *local* explanation methods are better suited.

One way to produce saliency maps for *local* explanations is by model-agnostic approaches (e.g., occlusion/perturbation-based methods; local interpretable model-agnostic explanation (LIME))^4,11–13^.They are model-agnostic, because such methods operate solely on the CNN’s input and monitor the produced output, making them unsuitable for explaining the CNN’s inner mechanisms^4^. To ameliorate this, attribution-based methods can be used^14^. However, there exists a plethora of model-specific attribution methods that are vastly inconsistent in terms of explainable outcome measures^15,16^, making it difficult for clinicians to ascertain which methods to select^4,8,10,16–22^. Moreover, these methods have mostly been used for *classification* (e.g., sleep apnea detection and EEG-stage classification using GCAM^23,24^, building XAI attribution ground truths for classification^13^, integrated gradients for diabetic retinopathy progression^25^), whereas the utility of XAI for *segmentation* – especially for medical imaging – has not been well investigated, which complicates the interpretation of attribution maps.

There are several challenges to meaningful interpretation of attribution maps. For example, CNNs may respond to very different structures than the specific anatomical markers prescribed by consensus guidelines typically used by clinical experts, who are not well equipped to read and interpret XAI signals. Regardless, attribution maps are often visually inspected, inviting bias and putting users at risk of missing critical information, further substantiating the need for automated methods. Accordingly, several works have indicated that XAI is promising but still requires more rigorous computational methods and analysis for reliable use^5,15,26^. Further, even for *classification*, the robustness of XAI methods to variations in training data is not well understood.

Here, we aim to take apart existing XAI methods and apply them in the context of auto-segmentation for radiotherapy, and to explore how these methods could be evaluated quantitatively. To this end, our contributions are four-fold. First, we take seven local, model-agnostic explanation methods commonly used for classification and adapt them to work for segmentation. Second, we compare all methods in an auto-segmentation setting using simulated data, where relevant parameters are controlled, and ground truths are fully known. Third, when the focus of each local explainability method is better understood, we evaluate each on clinical parotid gland (PG) contouring data. For this, we address the difference in XAI signal between (a) high- and low-performance cases, and (b) two different CNNs, both trained on clinical data but with varying manual reference consistency. Fourth, to use objective XAI as a tool to mark potentially inaccurate CNN-contours, we explore a topological data analysis approach for standardized interpretation of XAI signals. Combined, we provide relevant directives for clinical-use XAI and expose meaningful measures to work towards effective and safe clinical deployment of contouring CNNs.

## 2. Materials & Methods

### 2.1 Attribution-based XAI methods

This work used model-specific XAI methods: Guided BackPropagation (GBP), Gradient Class-Activation Mapping (GCAM), Guided GCAM (GGCAM), GCAM++, Layer-Wise Relevance Propagation (LRP) and PatternNet (PNET; signal + attributions). Although all methods share the same goal of providing the user with interpretable information of relevant input features, they result in different outputs inherent to their methodology. To help deploy them effectively for segmentation, we first review the most important aspects of each of their methodologies in the context of segmentation.

#### 2.1.1 Guided BackPropagation^18^

By combining the regular backpropagation algorithm (used during training) with deconvolution^10^ and a standard non-linear activation function (ReLU) on the backpropagated signal, guided backpropagation highlights positive contributions (features that activate neurons) and suppresses noise from negative contributions (features that deactivate neurons). This allows for fine-grained detail during visualization of important regions in an input image that contributed to the classification throughout all combined layers of the network. In the context of segmentation, this method can be repeated to accumulate contributions of multiple network output nodes for every predicted foreground voxel in the segmentation output.

#### 2.1.2 Gradient Class-Activation Mapping (Grad-CAM)^17^

Convolutional blocks used in CNNs behave as object localizers but lose this capability when the CNN is used for classification^27^. Class-Activation Mapping (CAM) is a method that can retrieve this localization information (saliency maps). CAM reconstructs this map *M_CAM_* by computing a weighted sum of n learned filters *F* in the final convolutional layer (Eq. 1):

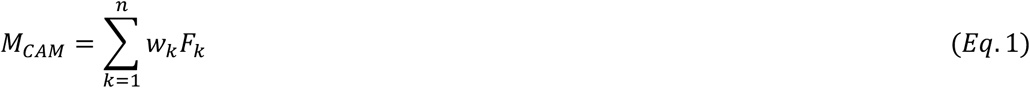

In CAM, logistic regression is used to estimate importance scores *w*. Instead, gradient CAM uses backpropagation to estimate w and construct the saliency map. GCAM has the advantage over GBP that it is class-discriminative (i.e. it can tell which activations are attributed to different classes), which is important for multi-class segmentation, and that contributions from any specific network layer with learned filters can be visualized. The main drawback is that it produces relatively noisy saliency maps by mapping entire filter maps, rather than contributions from individual neurons.

All GCAM variants can be used to obtain layer-specific information. For classification, the final convolutional layer is typically used, as it effectively presents a relevant summary of the input image, whereas this is not the case for segmentation^28^. Since there is no single layer in a UNet responsible for both encoding and decoding, instead, utilizing all layers from the decoder has recently been suggested for segmentation^29^. The last few network layers of GCAM are effective for classification because it maps spatial attributions of a single output value. In segmentation, this is not the case, and final layers mostly coincide with the predicted mask, which limiting GCAM’s relevance to segmentation. It has been suggested that compiling all bottleneck and decoder layers is more useful for segmentation^29^, because most of the localization information is processed there. Therefore, we follow this direction to obtain the attribution maps of GCAM and GCAM++, in addition to the final layer.

#### 2.1.3 Guided GCAM^17^

Guided GCAM is an adaptation of GCAM that combines the advantages of GBP and GCAM. Its attribution map, M_GGCAM_, is then obtained by computing the element-wise product of both (Eq. 2):

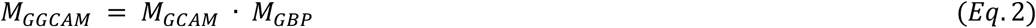

#### 2.1.4 GCAM++^30^

As a second adaptation of GCAM, GCAM++ was proposed to correct saliency maps for intra-class imbalances for e.g. occlusion of parts in the image normally needed for classification; thus, correcting for smaller structures compared to larger ones. GCAM++ achieves this by prioritizing contributions/voxels from smaller instances (i.e. increasing weights of voxels with smaller class instances during backpropagation), relative to other examples, leading to increased sensitivity. Similar to GCAM, the compiled decoder layers hold more meaningful for segmentation^29^.

#### 2.1.5 Layer-wise relevance propagation^20^

In a classification problem, the network assigns a certain classification probability before the final non-linear activation function. LRP is a method that goes backward through the network, to decompose to what extent each neuron in each previous layer was responsible for the output classification probability (i.e. relevance scores). This is repeated backward through the entire network, until voxel-wise relevance scores are obtained. Just like GBP, this produces fine-grained activation maps, but in addition, LRP allows increased flexibility by allowing use of different rule sets. In our implementation, a rule set that suppresses negative contributions is used^31^.

#### 2.1.6 PatternNet^32,33^

Although GBP and LRP reportedly carry interpretable information for CNNs, it was also observed that these gradient-based methods do not necessarily provide meaningful estimates of the XAI signal for any classification task^33^. This is because the noise present in an image may influence the CNN’s learning process^33^. PatternNet alleviates this issue by assuming that input features are a superposition of important characteristics of an image (relevant signal) and noise. The PNet signal (*S_w_*) is found by separating this noise from the relevant input features. Subsequently, PNet attribution (PNet-A) is obtained from the element-wise product of learned network weights with the denoised signal (Eq. 3):

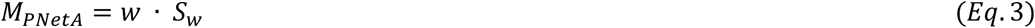

With this, the authors of PNet-A claim that it is an improvement over LRP, as it produces clearer individual voxel relevance score maps by disentangling signal from noise.

#### 2.1.7 Implementation

In this work, we directly apply m3d-cam^34^, an established PyTorch library for classification, as well as 2D and 3D segmentation networks, to contouring data to obtain saliency maps for GCAM and GCAM++. In addition, we use our own implementations for GBP, LRP and PNET, and adapt them to also work for 3D segmentation. This is achieved by repeating each respective method for every predicted foreground voxel in the segmentation output and subsequently accumulating and normalising the result. Since it was uncertain which GCAM layer was most useful for harnessing explainability, we visually compare the different GCAM layers. Finally, we implemented a manual multiplication of our own implementation of GBP and m3d-cam’s GCAM, to obtain GGCAM.

#### 2.1.8 Experimental workflow

The experimental overview (Fig. 1) of this work consists of (1) applying XAI to a synthetic setting where relevant parameters can be fully controlled, to understand the merits and limitations of each XAI method in the segmentation context; (2) applying and evaluating each method using (a) clinical, real-world radiation oncology data and (b) data with increased ground truth consistency, obtained by automatically removing cases with inadequate ground truths using a previously validated auto-curation method^35^; (3) qualitative evaluation of the synthetic and clinical context, and quantitative of clinical cases using topological data analysis.

**Figure 1.**
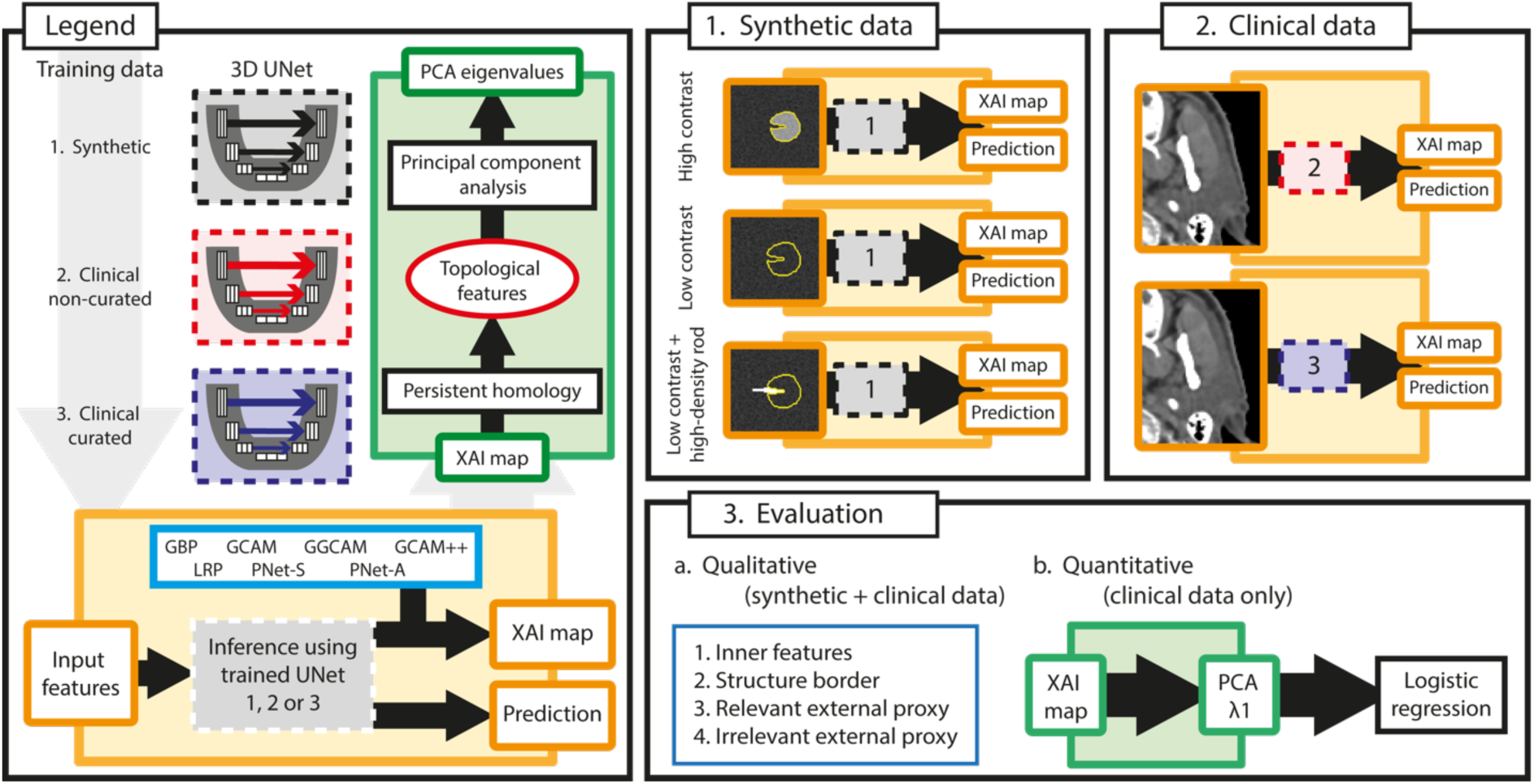
*Experimental overview.* Seven XAI methods were applied to a trained segmentation CNN in (**1**) a simulated setting using synthetic data; (**2**) clinical radiation oncology contouring data; (**3**) Evaluation then ensues through qualitative (simulated + clinical data) and quantitative (clinical only) measures.

### 2.2 Clinical background

The parotid gland is responsible for the production of saliva, has a high radiosensitivity^36^ and is one of the primary organs-at-risk (i.e. organs which are particularly susceptible to radiation damage and critical for preserving patient quality of life). Therefore, the parotid glands are among the most widely included structures to consider during treatment planning. Consequently, the limitations of parotid gland segmentations are relatively well understood, making the PG an attractive paradigm for investigating XAI for segmentation. Conventionally, PGs are manually segmented using computed tomography (CT) information. The PGs are located inferior to the lower half of the ear on the lateral surface of the cheek and extend to the lower border of the mandible^37^. PGs exhibit a large variety in terms of density, shape and size^29^, but generally consist of a larger superficial lobe, an inner lobe that folds medially (inward) around the mandible bone (Fig. 2; left), and occasionally an accessory gland (anterior lobe), located on the Stensen’s duct anterior to the main superficial PG lobe^37^. In addition to the shape and size variability, contouring can be challenging due to a patient-specific, low gray-scale contrast on CT^38^, and possible presence of dental artefacts, in which case XAI methods may be expected to use other structures as an (indirect) proxy for segmenting PG voxels (e.g. mandible bone).

**Figure 2.**
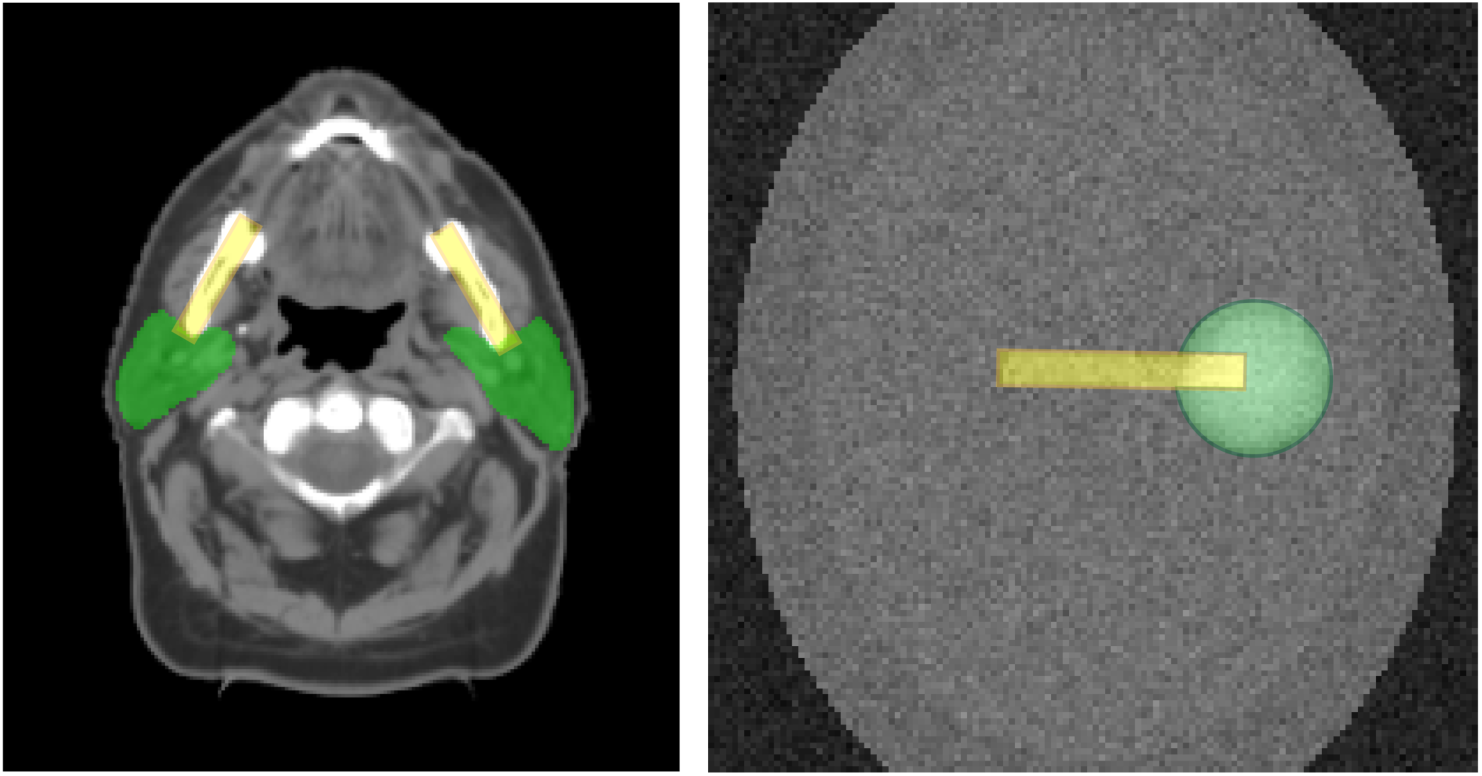
A real-world, clinical (left) and synthetic (right) situations of the parotid gland segmentation problems used to investigate XAI. The green contours in the clinical situation indicate both parotid glands, of which its simulated parallel is indicated by the green sphere. The mandible bone is simulated by the rod, indicated in yellow, pointing out of the parotid gland.

### 2.3 Synthetic parotid gland segmentation simulation

The robustness of XAI methods to contrast, training data and optimization stochasticity are not well understood. This complicates direct comparison of XAI methods in clinical settings because we cannot know whether a difference in XAI is inherent to the method or from external factors. In simulated/synthetic situations, we have full control over the CNN’s input, which we use to expose the CNN’s attributions with respect to structure contrast, and to investigate when it will start focusing on other, better visible structures (e.g. bone). This allows to make empirical sense of XAI outputs and to assess to which extent XAI maps agree to the clinical situation. Thereby, trust can be built that clinical XAI maps indeed also reflect meaningful information. To this end, we build a dataset where a ground truth is simulated^39^ in a simplified, simulated PG segmentation problem (Fig. 2). This is done by constructing a spherical “parotid gland” that folds around one end of a well-defined (simulated “mandible bone”) rod-structure, and then varying “PG” contrast.

Based on the densities of the constructed “parotid gland” (ρ_PG_) and “mandible bone” (ρ_MB_), six experimental variations (S1-S6) were defined (Fig. SF1). Each variation differed in 1) the (a) presence or (b) absence of the rod (ρ_MB_ = 1.0 if present or ρ_MB_ = 0.1 if absent), and 2) the density of the sphere, ρ_PG_. Values for ρ_PG_ were chosen such that (a) no information was present to detect it without the rod (i.e. density identical to s-HN); (b) it was barely distinguishable by the human eye (5% higher density than s-HN) but became indiscernible from s-HN after noise addition and (c) was clearly visible to the human eye (ρ_PG_ = 5 x ρ_HN_). Since the sphere was always attached to either one end of the rod, we hypothesize that the rod will act as a proxy for the CNN to identify the sphere, and that XAI methods should pick up on this proxy, especially when the sphere is poorly visible. For each variation, 300 cases were simulated, varying in the size of head-and-neck, the orientation and location of the sphere, and the orientation, location, and thickness of the rod. One CNN is trained for each variation using a train/val/test split of 240/30/30. Synthetic dataset creation details and an exemplary overview of synthetic variations are supplemented (SF1).

### 2.4 Data

CNNs for radiation oncology contouring are typically trained using data of variable quality^40^. It is, however, uncertain how sensitive XAI methods are to differences in training data and dataset quality. Therefore, in addition to a CNN that was trained with regular radiation oncology data, we also applied XAI methods to a CNN that was trained with an automatically curated training set, to minimize noise from imperfect contours present in radiation oncology contouring data and thereby to maximize interpretability of XAI results.

#### 2.4.1 Clinical data (a)

Our in-house clinical data used throughout this work consisted of 1925 PG contours paired with three-dimensional (3D) planning CT scans, acquired using a single GE HealthCare scanner. Scans and contours were collected as part of standard clinical protocols for a cohort of patients that were treated at our clinic for head-and-neck cancer between 2009 and 2021. The included dataset involved a typical head-and-neck case cohort, where no patients were pre-selected and no disease stages or sub-types were excluded. Contours were made according to consensus guidelines^41^ by 18 experienced radiation oncologists. Data pre-processing details are supplemented (SF5).

#### 2.4.2 Curated data (b)

Because it is currently unknown how robust XAI methods are to differences in CNN training data, we apply XAI methods using a CNN trained with more consistent reference segmentations, obtained through a previously validated approach^35^. To obtain the curated dataset, the regular clinical dataset was automatically cleaned by removing the 20% worst-performing training cases as rendered by the CNN early during training^35^. Here, the curation efficacy was defined as the DSC from curated networks minus non-curated network DSC.

### 2.5 Model training and validation

All models used throughout this work used identical hyper-parameter settings and 3D UNets^42^, which were adapted to use 4 depth layers, 32 input-layer filters (size 3×3×3 voxels), and a doubling of filters at each depth layer. This adapted network was used in a previously validated study^43^. Group normalization was applied after non-linear activation functions. Models were trained using four NVIDIA-GeForce GTX 2080Ti GPUs, with 11GB GPU RAM, 64GB system RAM and an Intel Core™ i9-9900KF CPU@3.6GHz processor. The GPU-version of PyTorch (Version 1.13) and Cuda 11.8, Python (Version 3.10.0) were used. Weights were initialized using Pytorch’s standard initialization method^44^ and models were trained using the ADAM optimizer^45^, using a combination of Dice similarity coefficient (DSC)^46^ and focal loss^47^ to guide model optimization (Eq. 4)^48^:

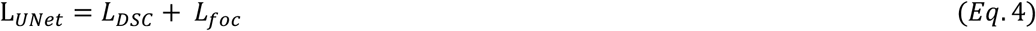

where focal loss mitigates the size-dependency of DSC loss and balances foreground and background voxels^44^. All models were trained using standard hyper-parameter settings, with loss hyper-parameters α = 0.5, ψ = 2.0, e = 1×10^−8^ and optimizer parameters β_1_ = 0.9, β_2_ = 0.999 and ε = 1×10^−7^, a batch size of 1 and an initial learning rate of 0.001. To mitigate divergence of model weights during training, Pytorch’s cosine annealing learning rate^49^ was used without warm restarts, with a minimum (final) learning rate of 1×10^−5^. For each experiment, the number of epochs required to fully train the network was heuristically chosen, using a train/val split of 1540/385 individual (uni-lateral) parotid glands. In the synthetic experiment, CNNs were trained for 25 epochs, which was the only difference in training settings compared to the clinical experiment. For curation experiments, since samples were removed, the number of epochs was adjusted such that an equal amount of data was seen by both the non-curated and curated CNNs, to ensure fair comparisons, and were optimized for 40 and 50 epochs, respectively. For every epoch, models were saved and validation loss was computed to determine the early stopping epoch after training. Data augmentations were performed on-the-fly, using Medical Open Network for AI (MONAI), an established, open-source, PyTorch framework^50^. Modes of augmentation were: (1) rotations (p: 0.2) of up to 15 degrees using bilinear and nearest-neighbour interpolations for CT and contours, respectively. (2) zooms (p: 0.2) between 0.9 and 1.1 scaling factor; (3) left/right flipping (p: 0.5); (4) intensity shifts (p: 0.2) with an offset of 0.1. Augmentations were turned off at test time.

### 2.6 Layer-specific GCAM explanations

Since the final convolutional layer has been the standard for selection of Grad-CAMs layers for classification, this has been included for analyses. However, as recently suggested by other works^29^, using decoders may be more suitable for segmentation, as these are responsible for the localization of important features in UNet. Therefore, a compiled and averaged map of decoder layers including bottleneck has also been included for analyses. Further, to better understand the local information used by the network across different levels of the network, all individual convolutional layers are displayed (Fig. 7). In addition, the GCAM attributions of the bottleneck + decoders, as well as all convolution layers in the network, are averaged and displayed (Fig. 7).

### 2.7 Evaluation

The American Association for Medical Physics (AAPM) head-and-neck segmentation challenge dataset from MICCAI 2015 was included as our external and independent evaluation set of 96 cases^51^. As a baseline performance reference, CNN contours are generated for all AAPM patients for both the regular and the curated network. Model performance was measured using DSC.

#### 2.7.1 Attribution map pre-processing

To ensure comparability between all methods, all XAI maps were processed identically as follows. To reduce the complexity of interpretations, negative contributions were nullified by subtracting the median (i.e. “neutral activation”) and casting all negative values to zero. Then, each attribution map was scaled between 0 and 1, where 1 corresponded to the maximum value for that specific map. The resulting maps are used for qualitative and topological assessment, and visualization.

#### 2.7.2 Qualitative saliency map assessment

There are several ways of evaluating XAI outcomes (e.g. localization/pointing game accuracy^52^, faithfulness^5,53^, explicitness^53^, stability^54^, fidelity^54^), but most of these metrics do not work for segmentation (e.g., pointing game relies on localizing relevant features within the predicted contour, which is not helpful for segmentation). In addition, XAI methods have been reported to depend highly on the specifics of the task at hand^15,16^, and because CNNs may attribute high importance to certain feature combinations / contrast subtleties that are irrelevant to humans and can quickly process much more data, humans are unsuitable for effectively interpreting CNN explainability. So, there currently exists no generally accepted way to evaluate XAI for segmentation, and we therefore base our evaluation on five empirical characteristics: The XAI highlights (1) voxels within the predicted foreground; (2) edges of the predicted foreground; (3) soft-tissue landmarks outside the predicted foreground in reasonable proximity to the PG (e.g. muscle edges); (4) high-intensity structures outside the predicted foreground in reasonable proximity to the PG (e.g. mandible bone, carotid artery, posterior skull base); (5) mundane structures, not deemed particularly important for PG segmentation (e.g. air, dental artefacts). Throughout all figures, empirical characteristics are indicated in figures using colors: (1: blue; 2: orange; 3: green; 4: pink; 5: black). For clinical data, XAI maps of 32 evaluation patients were scored (−−, −, 0, +, ++) across figures 3-6, where −− and ++ means low and high utility, respectively. For the final summation (Tab. 2), each score constituted −2, −1, 0, +1 and +2 points, respectively. The clinical scoring was done by three experts in the field of radiotherapy, each with 6, 12 and 22 years of experience, using a random subset of 1/3 of the AAPM evaluation dataset (=32 cases). The average of the three scores over all raters was reported.

**Figure 3:**
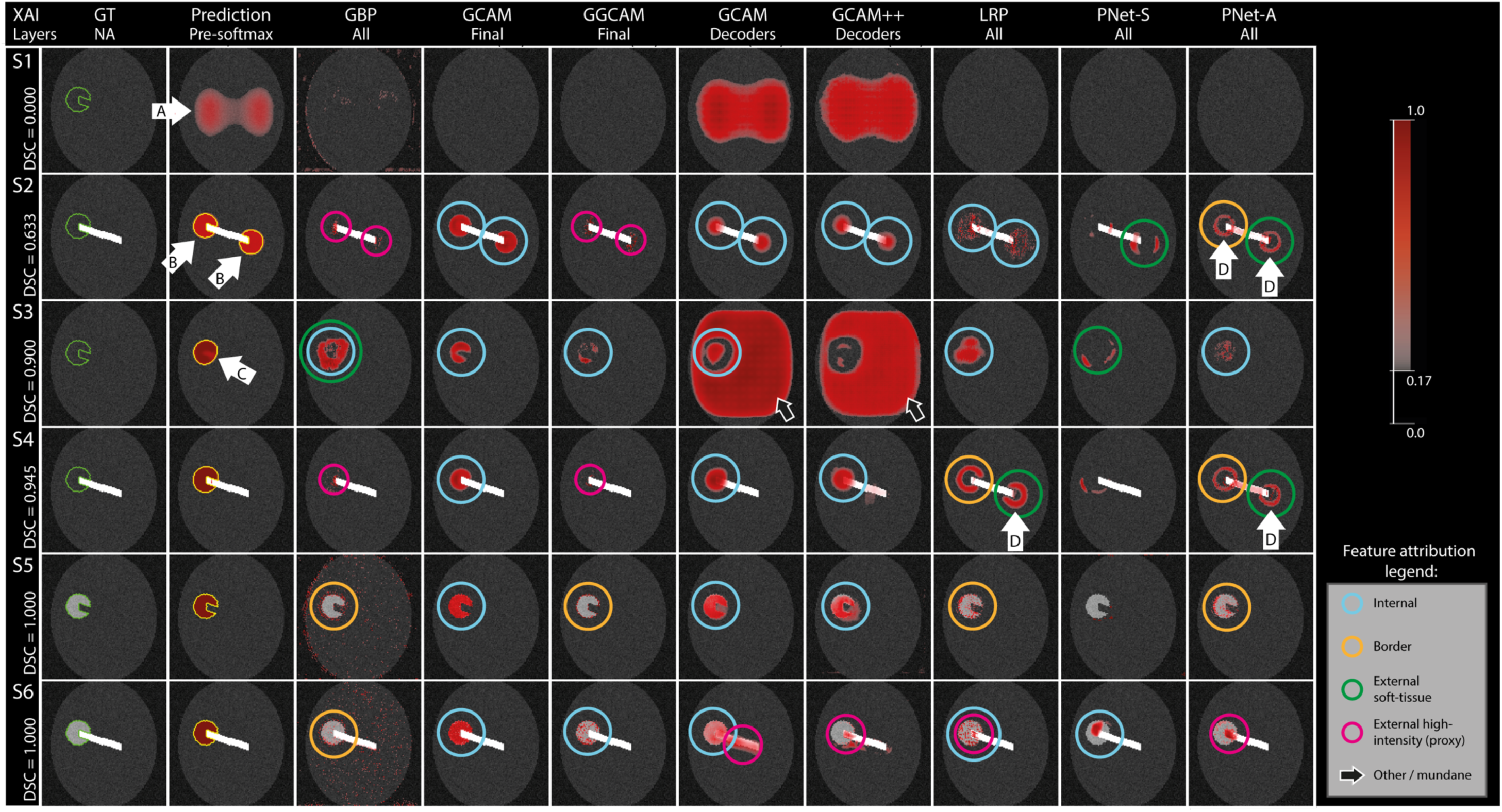
*Synthetic data XAI.* Contouring (GT: green contour; prediction: yellow contour) and XAI methods of synthetic test cases. DSC values indicate median performance. Arrows (**A**): CNN outputs the sphere’s probability density distribution. (**B**): CNN cannot distinguish the correct side of the sphere and therefore outputs both sides. (**C**): With low contrast, the CNN cannot identify the rod’s junction. (**D**): CNN implicitly learns the radius and shape of the sphere and attaches it to the rod. Abbreviations: GT: ground truth; GBP: guided backpropagation; GGCAM: guided gradient class-activation mapping; LRP: layer-wise relevance propagation; PNet-S: PatternNet signal; PNet-A: PNet attributions; DSC: Dice similarity coefficient.

#### 2.7.3 Topological data analysis

Model-specific attribution methods have been shown to be highly sensitive to the specific use-case and are vastly inconsistent in terms of outcome measures^3,18^. Moreover, metrics proposed for XAI evaluation do not work for segmentation (e.g. pointing game assumes the only relevant information resides within the contour), so a generally accepted way to evaluate attribution maps does not exist for segmentation. Moreover, as patient anatomies differ, attribution maps from different patients cannot be directly compared voxel-by-voxel for effective XAI analysis. Alternatively, XAI interpretation can be done visually, but humans are inherently unsuitable to process large amounts of data produced from the large amount of CNN parameters. In addition, this would introduce bias and put the user at risk of missing critical patterns. Therefore, more rigorous computational approaches are necessary for effective attribution map analysis^4^.

Persistent homology (PH) is a mathematical tool that algebraically derives topological signatures of complex numerical datasets and, while respecting anatomical differences between patients, can efficiently assess a large filtration range, which is difficult to judge visually. PH has been applied to various real-world problems^55^ (e.g. neuroscience^56,57^, cancer imaging^58^, radiomics for stereotactic body RT^59^). Such topological approaches can provide information about the underlying structure, shape and organization of data. More specifically, PH can be used to express level sets (i.e. equi-intensity maps) of typical XAI data across a range of thresholds in numbers of connected components (Betti-1), loops (Betti-2) and voids (Betti-3/Euler characteristic). GUDHI’s cubical complex^60^ was used to perform intensity filtrations of saliency maps for every AAPM patient. An eigen-decomposition (principal component analysis; PCA) was then performed on the resulting Euler characteristics, which embed information of each Betti-number. Thus, resulting eigenvalues represent the modes of largest topological variation. Eigenvalues were then fitted and correlated (Pearson’s, two-sided) with DSC to estimate the likelihood that the CNN outputs an accurate result. This method was performed for all XAI methods, using all available cases for evaluation in the AAPM dataset.

#### 2.7.4 Receiver Operating Characteristic

A relation between DSC and the first PCA eigenvalue was fitted by least squares minimization of a sigmoid with the resulting data points (logistic regression), with *A*, *k* and *λ_0_* as fittable parameters, and *λ* is the first PCA eigenvalue (Eq. 5):

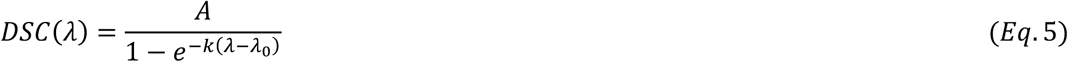

As proof of principle, to investigate whether this method can be used to identify the lowest DSC, we define an acceptance threshold of 15%, and determine the true positive and false negative rates of the classifier using this threshold. Subsequently, we use the area under receiver operating characteristic curve (AUROC) to report goodness-of-fit.

### 2.8 Visualization

To improve figure readability, a colormap was constructed that was transparent at low (<17%) signal levels and became brighter with higher signal levels. GCAM maps at deeper (pooled) levels, were up-sampled to match the original resolution. Since each XAI method requires a different approach for effective visualization, histograms of GBP, GGCAM, GGCAM++, LRP and PNet-A were adaptively equalized to aid visual assessment. For each method, an appropriate equalization cap was heuristically determined such that adequate visual maps were obtained.

## 3. Results

### 3.1 Simulated cases

#### 3.1.1 Segmentation

CNN segmentation performances drastically improved when the sphere was better visible (Tab. 1). Without sphere contrast and without the rod (S1), the CNN estimated the probability density distribution of the sphere, but could not predict its location (Fig. 3, arrow A; DSC = 0.0). When the rod was present, the CNN predicted spheres on both sides of the rod (Fig. 3, arrows B). With minimal sphere contrast and no rod, the CNN accurately segmented the sphere, but was inaccurate in the rod’s junction (Fig. 3, arrow C), whereas the presence of the rod further improved the segmentation. With high sphere contrast, the CNN achieved near-perfect DSC agreement with and without the rod.

**Table 1:** Agreement between the synthetic ground truth sphere and CNN model prediction. Displayed values are median[q1-q3] over AAPM data. Abbreviations: DSC: Dice similarity coefficient.

| Sphere | No contrast |  | Minimal contrast |  | High contrast |  |
| --- | --- | --- | --- | --- | --- | --- |
| Rod | *Absent* | *Present* | *Absent* | *Present* | *Absent* | *Present* |
| Variation | S1 | S2 | S3 | S4 | S5 | S6 |
| DSC | 0.0[0.0-0.0] | 0.63[0.62-0.65] | 0.91[0.90-0.92] | 0.95[0.94-0.96] | 1.0[1.0-1.0] | 1.0[1.0-1.0] |

#### 3.1.2 Explainability of synthetic segmentations

Examples of each method applied to synthetic data are shown (Fig. 3). Here, pink, orange and blue circles indicate highlights in the proxy (rod), sphere’s border and sphere inner structures, respectively. In all cases the final layer GCAM highlights coincided with the segmented volume but had lower activations than the segmentation before the final sigmoid activation function (Fig. 3; GCAM (fin). As there was no predicted mask when the sphere had no contrast and no rod present (Fig. 3, row S1), only decoder activations from GCAM and GCAM++ showed activations there. With no sphere contrast but with the rod present (Fig. 3, row S2), GBP and GGCAM highlighted the rod’s endpoints, which was the only relevant marker for accurate segmentation of the sphere (Fig. 3, pink circles). With low sphere contrast and no rod present (Fig. 3, row S3), GBP and LRP highlighted voxels globally across the inside of the sphere (Fig. 3, blue circles). With low sphere contrast and with the rod present (Fig. 3, row S4), GBP and GGCAM again showed activations in the rod’s endpoints (Fig. 3, pink circles), whereas LRP and PNet-A highlighted the sphere’s borders, and PNet-A showed activations both in the rod’s endpoints and the sphere’s edges (Fig. 3, orange circles). With high sphere contrast and without the rod (Fig. 3 row S5), GBP, GGCAM, LRP and PNet-A highlighted a few hyper-voxels along the sphere’s borders (Fig. 3, orange circles). With high sphere contrast and with the rod present (Fig. 3, row S6), all except GBP and GGCAM again highlighted few hyper-voxels along the sphere’s borders (Fig. 3, orange circles), whereas LRP, PNet-A and the decoder layers of GCAM and GCAM++ now highlight the rod. (Fig. 3, pink circles). Finally, PNet-A and LRP sometimes showed highlights of structures implicitly learned from the rod proxy (Fig. 3, arrow D).

### 3.2. Clinical cases

#### 3.2.1 Patient-specific explanations

By comparing multiple cases from the worst and best-performing DSC deciles, several trends were observed. First, GBP typically highlighted bony structures around the medial area of the PG (Fig. 4-6, GBP column) specifically of the mandible bone, contrast from the carotid artery and posterior skull base. This observation was consistent among different patients across the DSC distribution range (Fig. 3-6). Second, in high-scoring DSC cases, PNet-A showed more coherent activation patterns that also resembled the shape of a parotid gland (Fig. 4, C1-C6), compared to PNet-A in low-scoring DSC cases (Fig. 5, C2-C3, C5-C6). Additional examples are supplemented (SF. 4-5).

**Figure 4:**
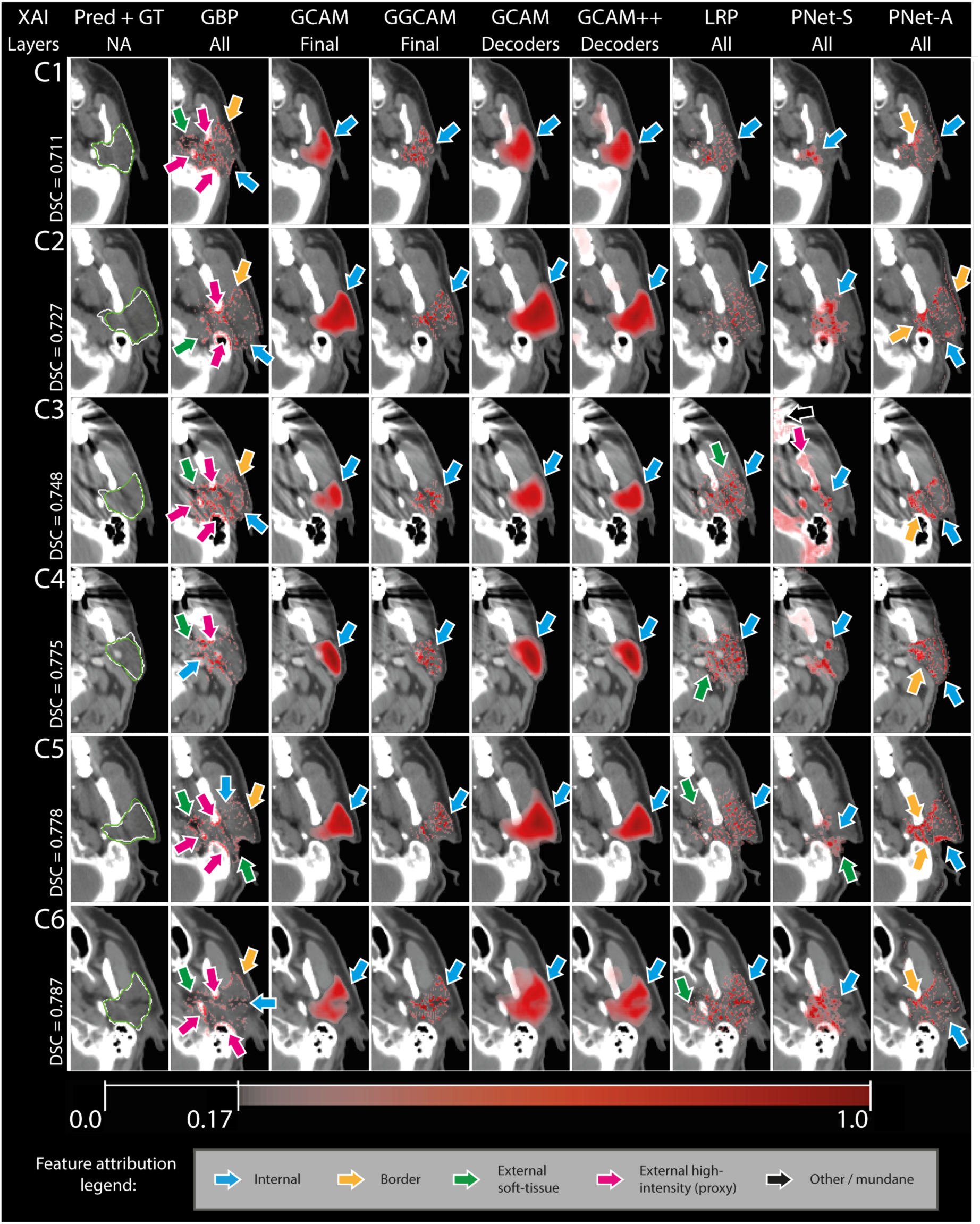
*High-performance clinical XAI.* Overview of XAI methods on clinical predictions comparing 6 patients randomly sampled from the 10% highest DSC decile using the curated CNN (C; green contour). Abbreviations: GBP: guided backpropagation; GCAM: gradient class-activation mapping; GGCAM: guided GCAM; LRP: layer-wise relevance propagation; PNet: PatternNet; PNet-S: PNet signal; PNet-A: PNet attributions.

**Figure 5:**
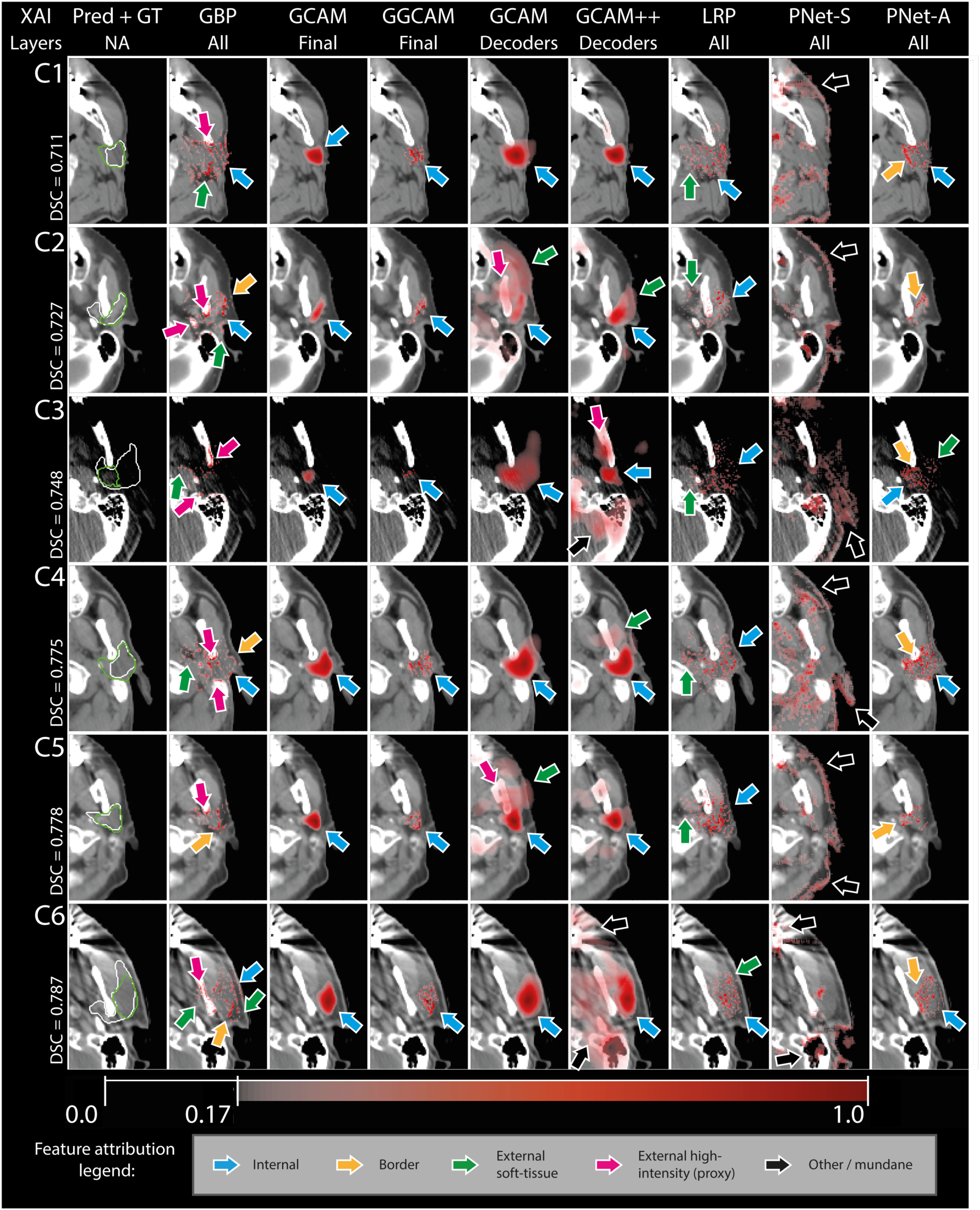
*Low-performance clinical XAI.* Overview of XAI methods on clinical predictions comparing 6 patients randomly sampled from the 10% lowest DSC decile using the curated CNN (C; green contour). Abbreviations: GBP: guided backpropagation; GCAM: gradient class-activation mapping; GGCAM: guided GCAM; LRP: layer-wise relevance propagation; PNet: PatternNet; PNet-S: PNet signal; PNet-A: PNet attributions.

#### 3.2.2 Model-dependent explanations

Non-curated and curated CNNs yielded median[q1-q3] DSCs in AAPM cases of 0.858[0.827-0.880] and 0.877[0.855-0.897], respectively. The observation that GBP highlighted bony structures around the medial area was more consistent in the curated network than in the non-curated network (Fig. 6, pink arrows). GBP, GGCAM, LRP, and PNet-A generally showed higher activations near lobes that were segmented more accurately (Fig. 6; anterior lobe: row C2; medial lobe: row C3). GCAM maps highlighted an area identical to the segmented volume (before the final sigmoid activation function), but with lower activations signal. GGCAM yielded higher detail of voxels within the prediction mask compared to GBP and GCAM. GCAM++ decoders did not deviate from GCAM decoders to a large degree.

**Figure 6:**
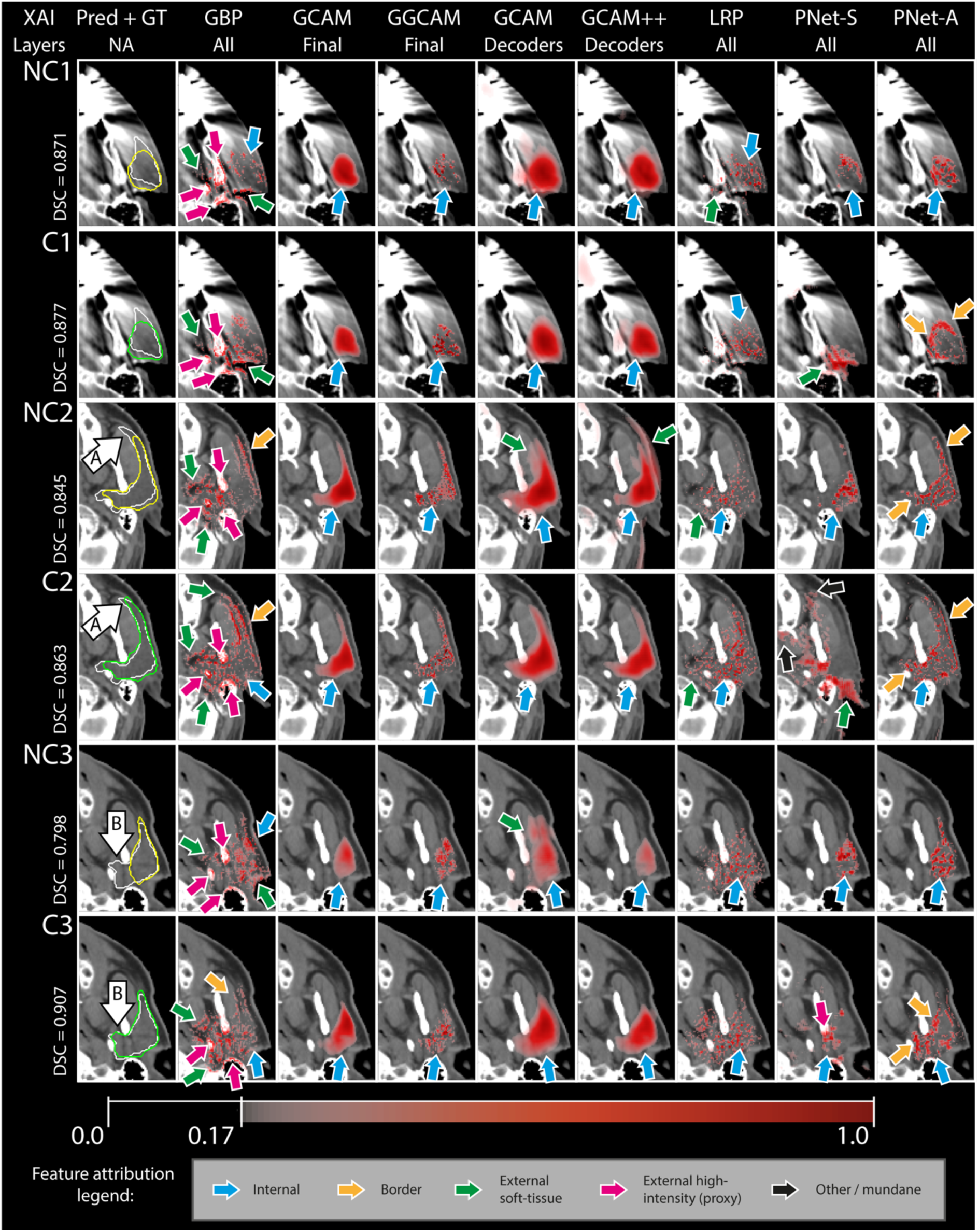
*Curated vs. non-curated model XAI.* Overview of XAI methods on clinical predictions comparing both non-curated (NC; yellow contour) and curated (C; green contour) CNNs. Arrows indicate notable sites of improvement from curation in anterior (A) and medial (B) lobes. Three patients (C1-C3) were randomly sampled from each tercile of the curated DSC distribution. Abbreviations: GBP: guided backpropagation; GCAM: gradient class-activation mapping; GGCAM: guided GCAM; LRP: layer-wise relevance propagation; PNet: PatternNet; PNet-S: PNet signal; PNet-A: PNet attributions.

#### 3.2.3 GCAM layer attributions

In general, the encoder pathway highlights contextual information of the CNN’s field-of-view. More specifically, layers earlier on in the network contain edge detectors (Fig. 7; E.1-1) and focus on high-density shapes (Fig. 7; E.1-2, 2, E.1-3, E.2-1, E.2-2). Then, activations are more abstract, but the CNN prioritizes soft-tissue details (Fig. 7; E.2-3, E.3-1, E.3-3, B.1-1) and localizes areas for which soft-tissue features were relevant in bottleneck layers (Fig. 7; B.12, B.1-3). The decoder combines these features to localization maps. The first decoder identifies an already rough version of the segmentation map, which is fine-tuned in the second and third decoder and final convolution layer. GCAM maps for the different layers were relatively consistent (i.e. highlight similar structures) across different samples.

**Fig. 7.**
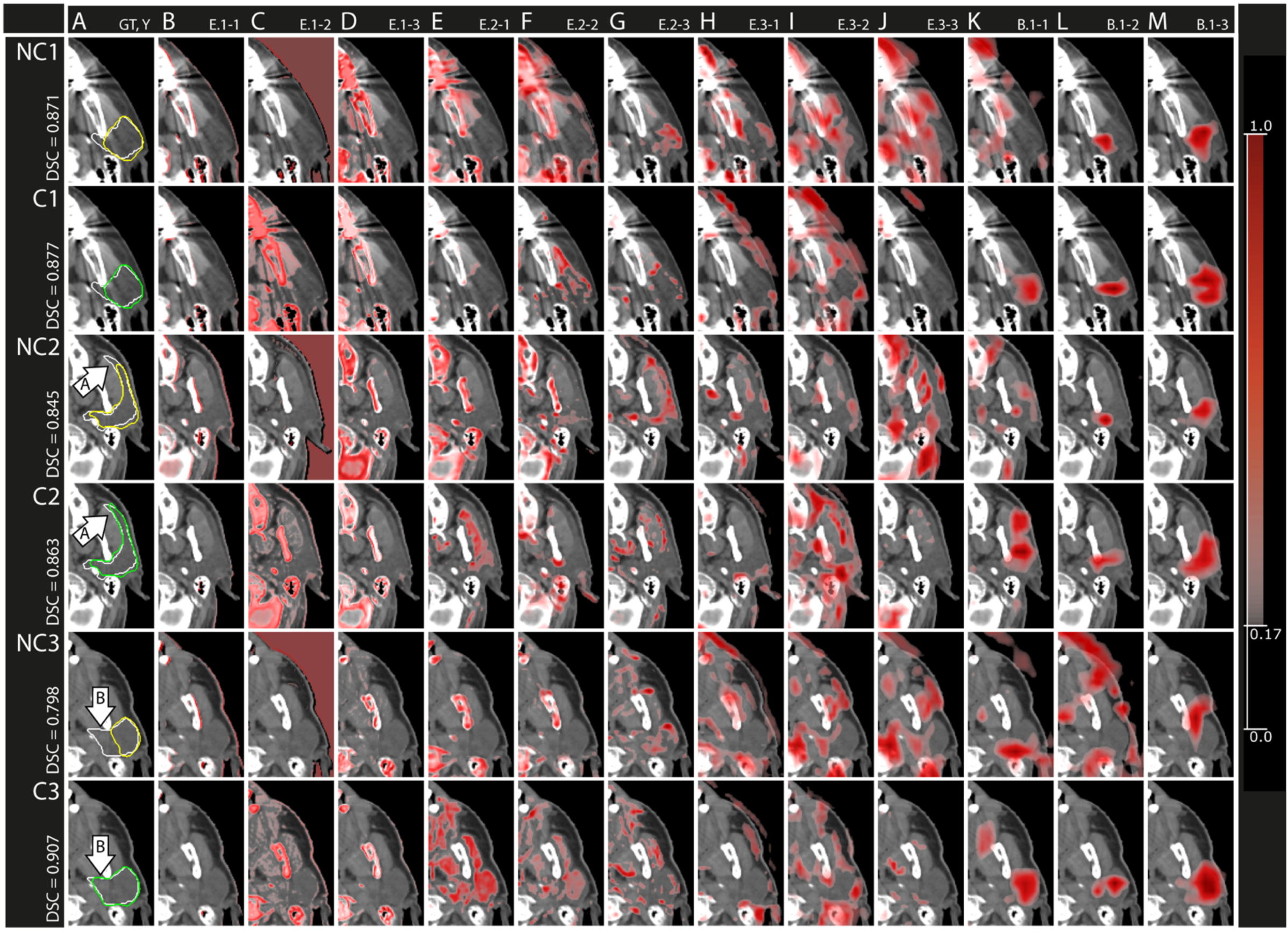
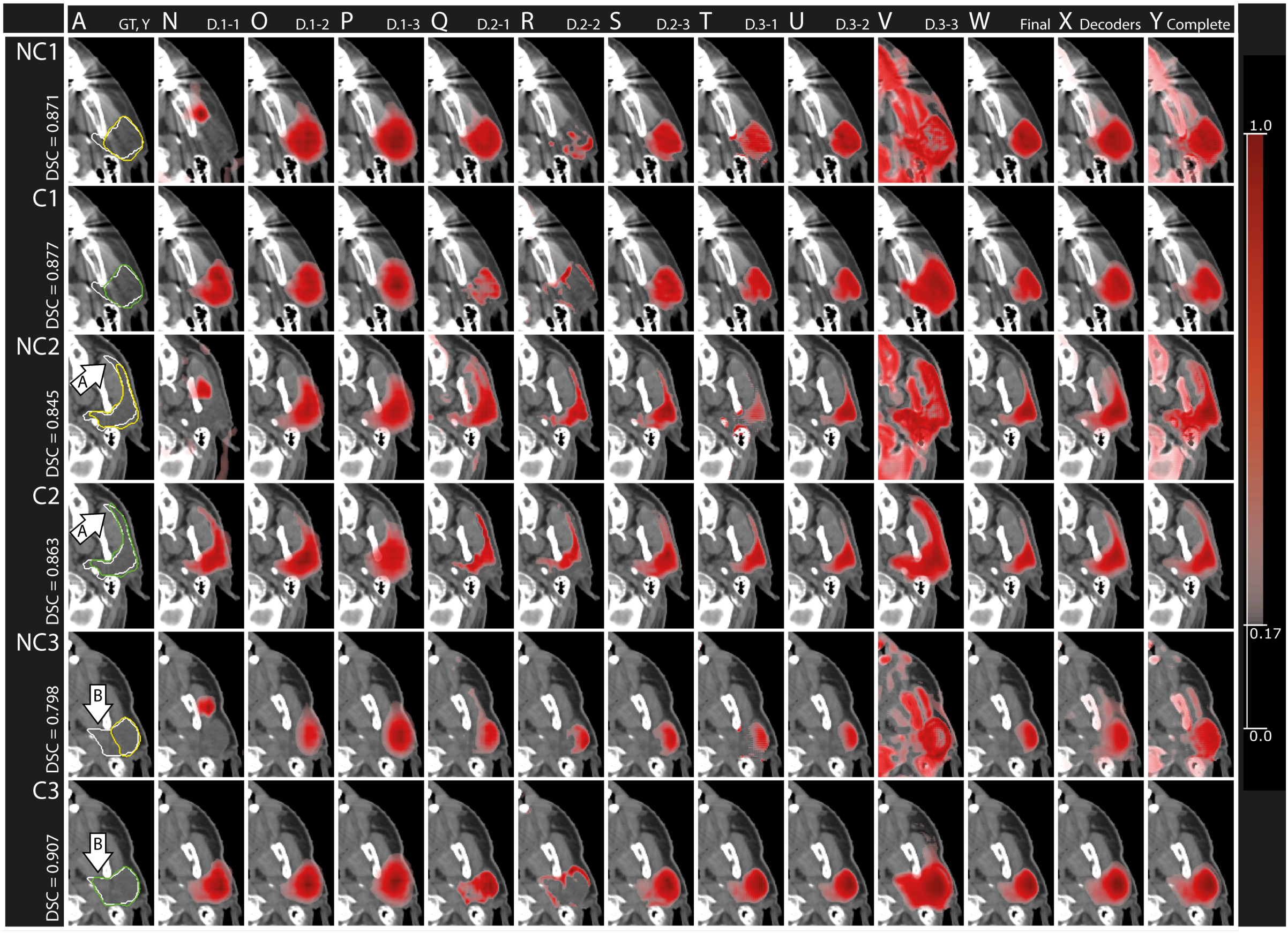
Cont. A*ttributions of individual GCAM layers.* Columns: E: Encoders; B: Bottleneck; D: Decoders, each of which contains three convolutional layers (1-3). Column A indicates the reference (GT; white) and predicted (yellow/green) contours. W denotes the network’s final convolution layer. X and Y show the average of decoder and full network layers, respectively. Odd and even rows contain results obtained using non-curated (NC; yellow) and curated (C; green) networks, respectively. Arrows indicate notable sites of improvement from curation in anterior (A) and medial (B) lobes. The three shown cases are randomly sampled from each tercile of the Dice similarity coefficient (DSC) distribution.

#### 3.2.4 Model-dependent GCAM layer attributions

GCAM of the curated CNN exposed different activations across virtually every layer along the encoder pathway, up to the last bottleneck layer. Specifically, in the first layer, non-curated CNNs highlight sharp horizontal contrast transitions across the image, whereas curated CNNs focuses more on contrasts in close proximity to the PG. Then, GCAM of the non-curated CNN showed higher activations in mundane features (i.e. air) or showed a more disorganized activation pattern, compared to the curated CNN (Fig. 7; rows NC3 and C3, columns C, H-L). Decoder pathway activations were more consistent in curated models but deviated substantially from its non-curated counterpart in the final convolution layer, which yielded cleaner activations in the curated model.

### 3.3 Persistent homology of saliency maps

The clearest trend between DSC and the first Euler characteristic PCA eigenvalue was found from GGCAM, with the highest AUC of 0.94, followed by PNet-A with AUC of 0.83. Other AUCs were 0.66 (GBP), 0.74 (GCAM), 0.62 (GCAM++), 0.62 (LRP) and 0.74 (PNet-S). PNet-A showed similar trends but was noisier. PH of GGCAM resulted in an approximated function that estimates a probability density distribution of CNN accuracy (Fig. 8a). With eigenvalues <-2.2e-3, our classifier flagged inconsistent contours with high (>85%) sensitivity and excellent (>90%) specificity. With eigenvalues >-1e3, ∼96% of contours belong to the highest 80^th^ percentile. We estimate that eigenvalues λ < −2.2e3, −2.2e3 ≤ λ < 0.3e3 and 0.3e3 ≤ λ respectively indicate high, moderate, and low risk for CNN contouring quality. The impact of two additional PCA modes are supplemented (Fig. SF3).

**Figure 8.**
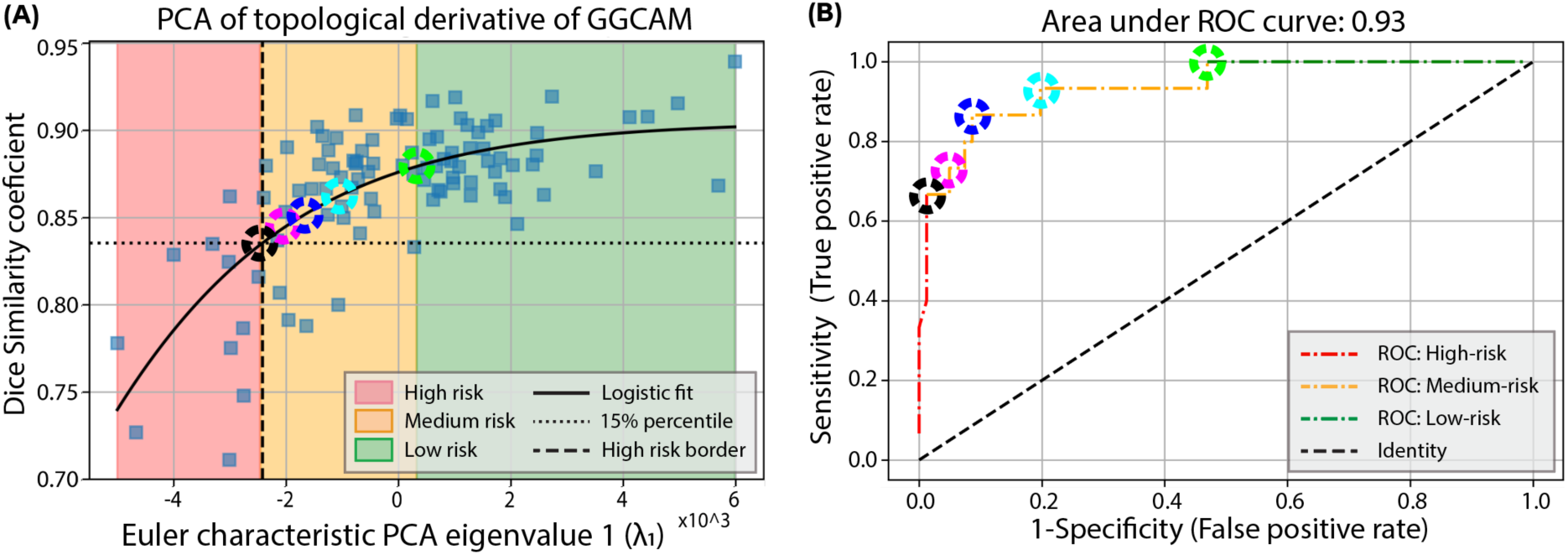
T*o*pological *derivatives of GGCAM signals of the complete AAPM dataset.* (**A**) PCA eigenvalues of Euler characteristic curves from intensity filtrations estimate the likelihood of inadequate (dashed red: cutoff) CNN contours for DSC. The black, pink, blue, cyan and green circles correspond to the points on the receiver operating characteristic curve in B. (**B**) Receiver operating characteristic curve for a binary classifier with an acceptance threshold at the 15% DSC percentile (black dotted line in (**A**)). The blue circle corresponds to an EC PCA eigenvalue of 1.8e-3 and shows a high sensitivity (>85%) and excellent specificity (>90%). Abbreviations: PCA: Principal component analysis.

## 4. Discussion

We demonstrated that patient-specific explanations could be obtained for CNN-based parotid gland segmentations using seven different XAI methods. Although XAI methods are all designed to highlight relevant information to the CNN, most methods disagreed in terms of important explainable parameters. GBP, followed by PNet-A generally agreed most closely with expert reasoning by highlighting directly (e.g. structure edges) and indirectly (e.g. low- and high-density structural proxies used for identifying segmentation borders) important voxels to determine which voxels belong to the parotid gland. Finally, this work is the first to demonstrate the feasibility of persistent homology for extracting topological derivatives of attribution maps to predict the accuracy of CNN-generated contours.

### 4.1 Simulated data

Simulations showed that CNNs are well (DSC>0.9) able to delineate structures that are completely indiscernible by the human eye (Fig. 3, row S3). In addition, since the main factor in which the sphere varied was its centroid location, the information required for near-perfect sphere segmentation is made up from the tips of the rod (which ends in the sphere’s center). GBP most consistently registers this landmark and GBP and LRP were similar when highlighting the sphere’s border. The largest difference between GBP and LRP arose when contrast was minimal (S3-S4). GBP still highlighted the rod when present, whereas LRP focused more on the sphere. This suggests that LRP is more likely to focus on hard-to-segment soft-tissue voxels, even when a clearly visible proxy is present, whereas GBP will only do this when there is no other indirect feature from which to derive the locations of the parotid gland. Contributions from structural proxies became irrelevant when the sphere contrast was sufficient. XAI methods were generally consistent in highlighting the sphere’s borders where the contrast was small (S3-S4). We believe that this may be due to CNNs attributing larger weights to neurons that segment more difficult-to-segment voxels.

### 4.2 XAI trends in clinical data

On clinical data, non-GCAM variants (except PNet-S) generally agreed by showing higher activation patterns in medial lobes and anterior lobes that were more accurately contoured (Fig. 6), especially in GBP and PNet-A. This could be because the curated network was trained using more consistent data, which better segmented these specific PG areas. PNet-A activations generally showed more disseminated patterns of intense activations when trained with less consistent data, whereas this pattern became more concentrated around the medial lobe, soft tissues around the mandible bone and the anterior lobe, when using curated models. This suggests that models highlighting segments that are typically difficult to segment may be more likely to perform well in these areas, and that higher activation patterns don’t necessarily relate to confident networks. However, correlating visual patterns with DSC should be done with care, as DSC is dependent on structure size, and because an “unconfident” CNN can still be correct. In case of small structures, a low DSC is unlikely to be reflected by the underlying XAI pattern (Fig. 5, C1, PNet-A). This was confirmed by PH of the GGCAM signal (Fig. 8), which essentially circumvents the size burden of predicted contours.

### 4.3 Topological data analysis for saliency map interpretation

We find that features within the predicted PG contour can be descriptive of the DSC resulting from this contour. This means that the likelihood that a resulting segmentation is adequate could be estimated using only features within the PG and that potential cases for CNN-failure can be marked. Considering that clinicians typically have only the predicted foreground to judge whether to trust a CNN segmentation, we believe this can be a useful methodology for clinical-use CNNs. With AUC=0.94, the obtained trend was comparable to other work that investigated model-agnostic approaches for classification^61^. We also find varying effectiveness of PH for different methods, where GGCAM showed the largest benefit. In particular, there is a discrepancy between qualitative effectiveness of GBP (Tab. 2) and the observed topological analysis. This may be partly explained by PH being more robust to signal noise than expected; GGCAM is a high-resolution explainability method that describes features only within the predicted mask, whereas other methods involve also external features. It could be that the current intensity-based PH filtration is not suitable to encapsulate essential features further away from the parotid gland. Regardless, since this is the first application of persistent homology to XAI analysis, we argue that PH for XAI holds potential and expect that future investigations of PH for XAI (e.g. segmentation borders, better standardized processing methods of XAI signals, negative XAI contributions, other XAI methods) can achieve even more convincing performance predictability for a wider range of applications.

**Table 2.** Utility indications from visual interpretations of XAI methods for the clinical segmentation problem. Indications 1-4 are considered with respect to the segmented foreground by the CNN. Possible values are −−, −, 0, +, ++, as rated on average in 32 patients by three expert observers.

| Method | GBP | GCAM | GGCAM | GCAM | GCAM++ | LRP | PNet-S | PNet-A |
| --- | --- | --- | --- | --- | --- | --- | --- | --- |
| Layer | NA | Final | Final | Decoders | Decoders | NA | NA | NA |
| (1) Inside | + | - | ++ | -- | + | + | 0 | ++ |
| (2) Border | + | -- | 0 | - | 0 | 0 | -- | ++ |
| (3) Outside low HU | ++ | -- | -- | -- | - | ++ | + | + |
| (4) Outside high HU | ++ | -- | -- | -- | - | -- | -- | -- |
| (5) Other | ++ | 0 | ++ | + | - | ++ | -- | ++ |
| **Total (average):** | 8 | -7 | 0 | -6 | -2 | 3 | -5 | 5 |

### 4.4 Relation between simulations and clinical XAI

Several of the findings in simulated data were observed to extrapolate to clinical data. First, GBP showed activations in the bony structures (Fig. 4-7, pink), confirming our hypothesis that XAI registers easily observed markers for PG segmentation, especially without clear contrast of the segmented structure. Second, the same difference between GBP and LRP in simulated data (that LRP stops focusing on high-contrast proxies when the segmented volume is not easily discernible) was observed in clinical data, as the mandible bone was generally not highlighted by LRP. Finally, since PNet was designed to deconvolve relevant feature signals from noise, PNet-A should in theory produce a cleaner signal than LRP^34^. Although this was not directly clear from our observations (LRP showed lower activations), this indicates that PNet-A better picks up on subtle but important features than LRP.

### 4.5 GCAM layers

Considering the consistency of GCAM layer activations over multiple cases, the activations are robust to inter-sample differences. However, when comparing both (non-)curated CNNs, layers respond to very different structures up to the last bottleneck block (Fig. 7; rows NC-C). Moreover, it was observed that curated CNNs generally highlighted better organized (i.e. concentrated high-intensity) patterns in GCAM (Fig. 7; col. V). This may be an indication that bias reduction by curation may indeed further stabilize CNN training. From visual assessment of GCAM and GCAM++ signals across all layers, it remains unclear how GCAM can be useful for segmentation purposes and we would argue that its greatest utility is for classification. However, GCAM and its variants are indirectly useful to segmentation through persistent homology of the GGCAM signal.

### 4.6 Clinical-use XAI recommendations for OAR segmentation

From visual interpretations of both simulated and clinical XAI signals, we conclude that GBP, provided the most concise yet reliable explanations of CNNs for PG segmentation (Tab. 2), followed by PNet-A. We believe that these methods can harness relevant information of meaningful and anatomical markers to judge important features used for CNN-based contouring. This could potentially be useful (1) to judge whether a network was adequately trained (Fig. 6-7) and (2) as an indication that difficult-to-segment areas in OARs (e.g. medial, anterior PG lobe) have been picked up by the CNN. For example, for higher-scoring DSC cases, PNet-A exhibited more concentrated activations around PG edges and medial/anterior lobes, and more diffuse (i.e. more widely distributed) activations for lower DSC cases. This observation was in line with simulations, where reference segmentations were fully known and segmentations are less difficult for the network (Fig. 3; S5-S6, PNet-A). In addition, a better organized pattern in GCAM layers (especially the last convolution before the final segmentation layer) could be used to confirm stable training for PG segmentation. It should be mentioned that LRP showed promising performance in synthetic data, but this was not observed in clinical data. However, since XAI methods can be highly situational^15,16^, this does not necessarily mean that LRP cannot harness any meaningful explanations. Therefore, we suggest that LRP be included in additional work investigating the utility of LRP for segmentation. Further, GBP consistently highlights the mandible, carotid artery and posterior skull base for accurate segmentation of medial lobes, both for high- and low-DSC cases. This observation was less outspoken in the non-curated model. Taken together, we suggest using GBP to expose important anatomical markers for structure segmentation, but to complement it with PNet-A to determine more intricate attributions.

### 4.7 Limitations

First, most of this work relies on subjective, qualitative observations. Although we tried to objectify our interpretations as much as possible, unbiased interpretations cannot be guaranteed. For example, the choice of how XAI maps were normalized directly influences visible activations. This could cause potentially important information to be missed when interpreting XAI maps. Therefore, to mitigate this limitation, we also investigated a proof-of-concept topological approach to saliency map analysis. Second, in simulated experiments, the sphere was not designed to contain any features inside its contour that would aid its segmentation. This was done to maximize the interpretability of XAI results by controlling any potentially confounding variables. However, since GCAM and GGCAM focus on features within the segmented foreground, the full potential of GGCAM could not be properly harnessed using our simulated experimental setting. Considering the relatively successful interpretation of GGCAM using persistent homology, this was not likely to be an issue in clinical data. Third, the AAPM data used for XAI evaluations was from a different distribution of patients, which could potentially affect model explanations and should ideally have been complemented by in-distribution but clean datasets. Obtaining this dataset a difficult endeavor that risks introducing additional confounders, further complicating interpretation of XAI results, which is why we chose not to do this.

### 4.8 Future recommendations

Several recommendations for future work ensue from this study. First, while GBP showed considerable success in terms of qualitative assessment (Tab. 2), this was not confirmed by persistent homology approach (AUC=0.65). GGCAM generally exhibits the cleanest attribution maps, so it can be that persistent homology is sensitive signal noise. This could be mitigated by involving a geometrical persistence filtration. Second, considering GBP’s strong capability to highlight structural proxies not in or directly near the segmented volume, we expect GBP to be useful for explaining segmentation tasks that use anatomical landmarks for their segmentation (e.g. head-and-neck elective target, lung gross target volume)^62,63^. We hope that our work, which addressed XAI for the parotid gland as a well-understood segmentation paradigm, could be used as a steppingstone for future studies to investigate whether GBP confirms these anatomical landmarks in radiotherapy target volume segmentation. Third, segmentation was practically handled as repeated localization and classification of voxels, assuming equal importance for all voxels. However, since segmentation is essentially identifying border contours of a structure, it could be argued that XAI signals from border voxels should be prioritized over voxels located more inside the predicted contour. Future studies could consider obtaining XAI signals from only a selection of foreground voxels, such as structure borders or areas known to be more difficult to segment, such as the medial and anterior parotid gland lobes. Third, we only included DSC for evaluations and for defining our binary classifier. This is an arbitrary choice that was made for proof-of-principle exploration of PH as a potential tool for XAI analysis. Since DSC is limited when addressing clinical significance for radiation oncology purposes, future work should include distance metrics to better investigate the clinical relevance of PH. Fourth, we excluded negative contributions (i.e. features that make foreground segmentation less likely) from saliency maps. Accordingly, future studies could investigate whether confounding features for segmentation can be found, or whether including negative attributions’ persistent homology may further improve DSC predictability. Finally, we observed varying effectiveness of PH for different XAI methods. To mitigate this, future studies could consider a geometrical filtration or a combination of intensity + geometrical filtrations could be considered, or GBP and PNet-A could be combined to harness the advantages of both soft-tissue and high-density attributions. Finally, the list of XAI methods may be extended with local, model-specific methods suggested by Captum^64^, e.g. integrated gradients^22^, SHAP-methods^65,66^ and DeepLIFT^67^.

### 4.9 Conclusions

A lack of intuitive explanations of CNNs for segmentation still limits effective implementation of deep learning in the clinical workflow. This work showed that from seven common XAI methods, Guided BackPropagation and PatternNet attributions subjectively agree most with human expert reasoning on directly (e.g. segmentation border) and indirectly (e.g. structural proxies used for identifying segmentation borders) important features for parotid gland segmentation. However, XAI methods are still inconsistent in terms of explainable features and require more rigorous computational methods for analysis. This work is the first to show conceptual proof for the feasibility of topological data analysis of as an objective and potentially clinically useful quality assurance tool for segmentation convolutional neural networks. Although it does not directly clarify explainable features, topological data analysis may be used to derive confidence indices for generated contours.

## Data Availability

Our medical ethics committee has not given permission to share data due to reasons of privacy protection.

https://www.aapm.org/GrandChallenge/GDP-HMM/

## Data and code availability statement

Due to patient privacy, the data used in this study is not made publicly available. Code regarding preprocessing, XAI methods and persistent homology analyses are made available.

## Ethics statement

Due to the retrospective nature of this study, not including tests or different treatments on human individuals and the use of pseudonymized data, it was concluded by the Medical ethics committee that the Medical Research involving Human Subjects Act (WMO) does not apply to this study and that official approval of this study by the committee is not required.

## Vitae

Victor Strijbis MSc is a developing professional and biomedical engineering PhD student at Amsterdam UMC, with artificial intelligence, radiotherapy, medical imaging and computational physics as his main areas of expertise. His work focuses on the development and implementation of fast and adaptive engineering solutions for precision radiation oncology.

Dr. Oliver Gurney-Champion is a prominent researcher in medical image acquisition and analysis, specializing in advanced quantitative MRI techniques for precision medicine. Dr. Gurney-Champion focuses on the integration of AI and machine learning techniques into clinical imaging protocols. His work, widely published in top-tier journals, aims to enhance the precision and efficacy of oncological imaging, ultimately improving patient outcomes.

Dragos Grama MSc is a developing professional and machine learning engineer, with AI, computer science and radiotherapy as the main components of his portfolio. His works focus on the development and application of novel AI techniques to automate treatment planning in radiotherapy.

Berend Slotman is Professor of Radiation Oncology at Amsterdam UMC 1998 and was chair of this department for 28 years. Dr. Slotman received both his MD and PhD with highest honors at VU University in Amsterdam and was registered as radiation oncologist since 1994. He (co-)authored around 500 peer-reviewed publications and has coordinated several trials, including the EORTC study on prophylactic cranial irradiation in extensive stage small cell lung cancer. He is currently President of the Radiosurgery Society, and past-president of the American Radium Society and European SocieTy for Radiation Oncology.

Wilko Verbakel (PhD, PDEng) is an associate professor and senior medical physicist working at the radiation oncology department of the Amsterdam UMC, since 2004, and a director of research science at Varian, since 2023. His research in radiotherapy involves automated treatment planning, development of knowledge-based planning, implementation of lung and spine SBRT, tumor tracking, deep learning in radiation oncology, adaptive radiotherapy and FLASH treatment planning. Verbakel has over 130 scientific publications in international peer-reviewed journals. He has an interest in making optimal, efficient and advanced radiotherapy available all around the world.

## CRediT author statements

**Victor Strijbis:** Conceptualization, data curation, formal analysis, investigation, methodology, software, validation, visualization, writing – original draft, writing – review and editing; **Oliver Gurney-Champion:** Conceptualization, methodology, supervision, writing – review and editing; **Dragos Grama:** Data curation, software, writing – review and editing; **Berend Slotman:** Project administration, resources, supervision, writing – review and editing; **Wilko Verbakel:** Conceptualization, data curation, methodology, project administration, resources, supervision, writing – review and editing.

