## Supplementary Material A for "Interpreting convolutional neural network explainability for head-and-neck cancer radiotherapy organ-at-risk segmentation"

### Supplementary Material A (Supplementary methods)

#### Synthetic case generation

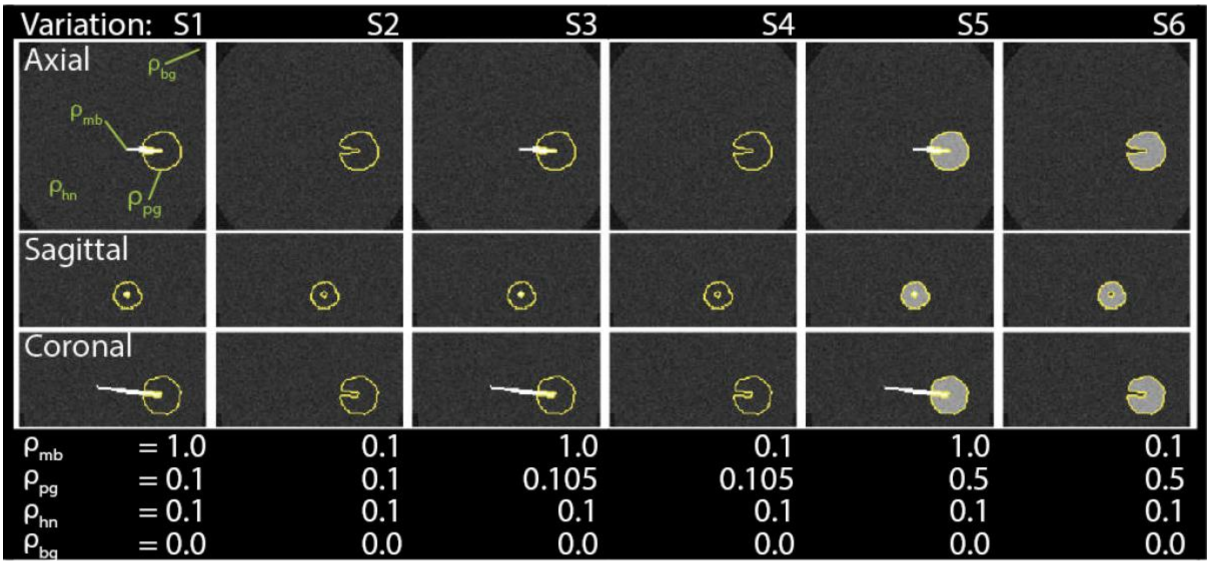


**Figure SF1:** Overview of synthetic experiment series S1-S6 for one virtual case, with increasing visibility of the PG, either with or without a visible stick (MB). $\rho_{BG}$, $\rho_{HN}$, $\rho_{MB}$, and $\rho_{PG}$ denote the background, s-HN, s-MB and s-PG densities, respectively. The yellow contour indicates the constructed ground truth.

The data for synthetic experiments (Fig. SF1) was generated as follows. A three-dimensional (3D) distance matrix of size 256 × 256 × 128 was constructed, where the voxel values indicated their minimal distance to the Z-axis centerline. This distance was scaled using horizontal and vertical spacing scalars $r_1$ and $r_2$, respectively, to construct a large, oval-shaped cylinder, and a smaller one placed on top of it, roughly simulating the head. We then constructed the "head"-tissue cylinder mask H by simulating the head size using scalar $H_R$. The top and bottom of H were indicated by Z-coordinates $H_{Z1}$ and $H_{Z2}$. Subsequently, the second "neck" cylinder N with radii 80% of the radii in C was attached at the bottom of C, forming a synthetic "head-and-neck" (s-HN) tissue-complex, in an "air" background. Each synthetic structure (background, head-and-neck and mandible bone) were given virtual densities of $\rho_{BG}$, $\rho_{HN}$, $\rho_{MB}$ = 0.0, 0.1, 1.0, respectively. We then simulated a sphere-rod complex, which resembled a parotid gland (s-PG) folding around the mandible bone (s-MB). Here, the rod had a virtual density $\rho_{MB}$ of 1.0, and the s-PG virtual density $\rho_{PG}$ depended on the experiment and was either equal to $\rho_{HN}$ (= 0.1), 0.105 or 0.5. The sphere radius $r_{PG}$ = 13 voxels was fixed to maximize interpretability of XAI signals. The rod was attached horizontally, originating from the centre of the sphere and had length $B_L$ and height/width $B_{WL}$. The rod-sphere complex was shifted by $t_x$, $t_y$ and $t_z$. Then, Gaussian noise ($\mu = \rho_{HN}$, $\sigma$ = 30% × $\rho_{HN}$) was subsequently added and voxel intensities were window-levelled between 0 and 1. Finally, for memory considerations, and because networks typically do not require the entire head-and-neck volume for adequate performance, FOVs were made by selecting the middle 128 × 128 × 64 voxels in each dimension. A summary of the parameters used to effectuate training variation is shown in Supplementary Table A.

| Parameter | Meaning | Domain | Distribution | Localization (μ) | Spread (σ) |
|---|---|---|---|---|---|
| $r_1$ | Horizontal grid scalar | [0.6,0.8] | Gaussian | 0.7 | 0.05 |
| $r_2$ | Vertical grid scalar | [0.8,0.9] | Gaussian | 0.8 | 0.025 |
| $H_R$ | Head size scalar | [80,110] | Uniform | 95 | 8.7 |
| $H_{Z1}$ | H caudal end | [32,42] | Uniform | 37 | 2.9 |
| $H_{Z2}$ | H cranial end | [112,120] | Uniform | 117 | 2.3 |
| $\alpha$ | *Inclination (XY-angle)* | *[-30,+30]* | *Gaussian* | *0* | *30* |
| $\theta$ | *Azimuth (XZ-angle)* | *[-30,+30]* | *Gaussian* | *0* | *30* |
| $\phi$ | *Zenith (YZ-angle)* | *[-30,+30]* | *Gaussian* | *0* | *30* |
| $t_x$ | X-translation | [-10,+10] | Uniform | 0 | 5.8 |
| $t_y$ | Y-translation | [-15,+15] | Uniform | 0 | 8.7 |
| $t_z$ | Z-translation | [-10,+10] | Uniform | 0 | 5.8 |
| $B_L$ | Rod length | [10,20] | Uniform | 15 | 2.9 |
| $B_{HW}$ | Rod height & width | [1,4] | Uniform | 2.5 | 0.9 |

**Supplementary ST1:** Tabular overview of the parameters that were variable for each synthetic example. Localization μ and spread σ indicated the centre and standard deviation of the corresponding sampling distributions, where σ of the uniform distribution was given by sqrt((b-a)$^2$/12), where a and b are the lower and upper domain limits, respectively.

**Preliminary experiments**

To determine an appropriate training epoch number for optimizations in simulations and clinical data, training and validation losses were monitored during the optimization of 5 models with different initializations for each experiment line. The point of convergence was defined as the point where the mean validation loss over all models plateaued. We found that this point happened only at very late stages of training, suggesting that the performed augmentations and learning rates were effective in preventing overfit. The training epoch was chosen as the epoch where, on average, models achieved maximal validation performance, while also minimizing training time. In practice, this point was reached at 25 epochs in simulated data and 40 epochs in clinical data.

**Persistent homology**

The persistent homology (PH) analysis relies on several pre-processing settings and choices specific to PH. Here, these choices are summarized. First, since PH requires computing topological profiles between all voxels for every step along the filtration, computation time and memory use increase exponentially with increasing included features. Therefore, we minimized the features to consider by defining a bounding box. The dimensions of this box were variable and based on thresholded XAI attribution values (median + std) (Fig. SF8).

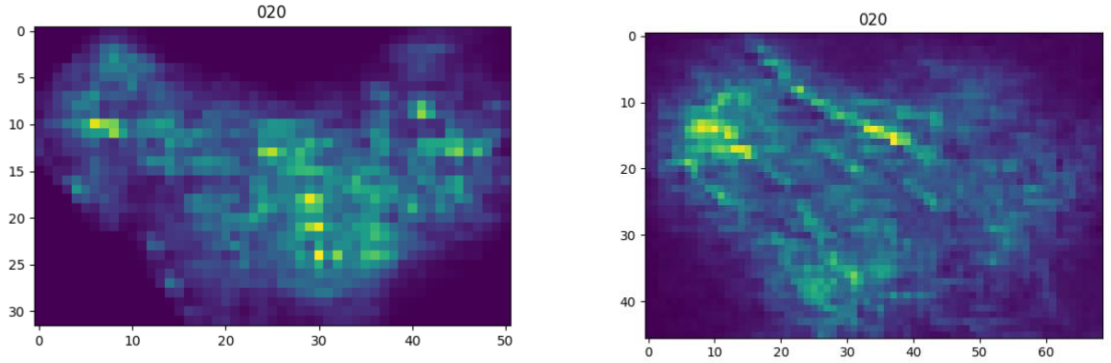


**Figure SF2** Two random example of bounding-boxed inputs of XAI attributions of the parotid gland (left: GGCAM; right: GBP) used for the filtration. Input was obtained using an attribution threshold (after normalization) of 0.01. The displayed 2D image is a projection (i.e. averaged over all slices) of the 3D input, but the original 3D input was used for the filtration.

Then, we used GUDHI's cubical complex to efficiently perform a filtration from the scalar field (intensity filtration of the bounded mage), which captures how connected components, loops and voids (Betti-numbers 0, 1, 2) form and disappear, when connecting voxels from varying grayscale intensities (i.e. level-sets). This intensity range over which these topological features persist is registered in the barcode plots (Fig SF9). Then, the barcodes of each respective Betti-number are summed for every increment on the x-axis, resulting in three Betti-profiles and an Euler characteristic profile ($Betti_0$-$Betti_1$+$Betti_2$) (Fig. SF10). These profiles were collected for every patient, and a PCA was performed where every bin along the x-axis was one feature, after which the resulting eigenvalues are correlated with DSC.

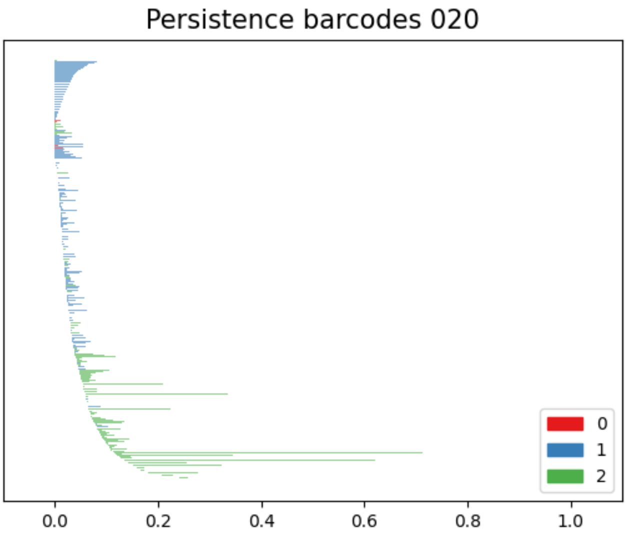


**Figure SF3** Barcode plot denoting the birth and death of connected components (red; 0), circles (blue; 1) and voids (green; 2). Visualisation is done using GUDHI's function plot_persistence_barcode.

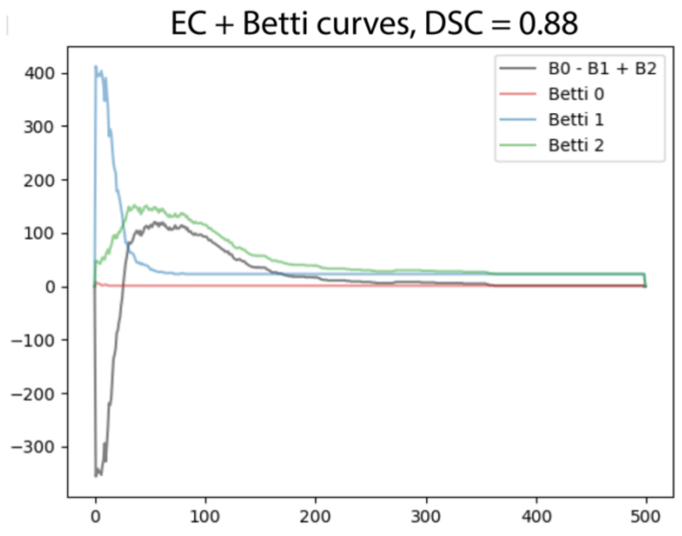


**Figure SF4** Example Betti and Euler profiles.
