## Supplementary Material B for "Interpreting convolutional neural network explainability for head-and-neck cancer radiotherapy organ-at-risk segmentation"

### Supplementary material B (Data acquisition and pre-processing)

In-plane acquisition resolutions were (0.80-1.27)mm, with a median[interquartile range (IQR)] of 0.98[0.97-1.02]. The slice thickness was 2.5mm in 96% of cases and 1.25 mm for the remaining cases. In-plane acquisition dimensions were 512x512x(98-581). All patient planning CTs and annotations were interpolated to a 1x1x1.25mm$^3$ voxel spacing by 3$^{rd}$-order spline and nearest-neighbor interpolation, respectively. This spacing was chosen to as an optimal trade-off for resolution (to allow for better erosions in corruption simulation experiments) while minimizing interpolations. To accommodate dose calculation, radiotherapy CTs contain a large field-of-view (FOV). To reduce the memory required for our networks, we wrote an automated algorithm that cropped the CT (176x192x176 voxels) to only contain the relevant regions of the CT necessary for segmentation. As patients were placed in the scanner with varying native positions and neck flexions during treatment, patients' heads showed large positional variations in CT DICOMs. Using the same image coordinates for constructing the field-of-view (FOV) for all patients would cause parotid gland to fall outside of it for some. We ensured that for all patients, both parotid glands were encompassed in the FOV by algorithmically finding the 6 borders for each patient individually (Fig. SF1.1). First, the couch was identified by selecting the axial slice where the derivative of the slice-averaged Hounsfield Units (Hus) exceeded one standard deviation (Fig. SF1.1a; red arrow), after which it was removed from the planning CT by casting all couch slices to air HUs (-1000). Second, the cranial end slice of the FOV was detected. To do this, the CT was first window-leveled to between -200 and 300, which was previously reported to serve as a good window leveling for most head and neck automated segmentation tasks[68]. Then, a cranial reference point was detected as the point in the face area where the HUs summed along the axial axis was maximal (Fig. SF1.1b; blue arrow). From this, the cranial border was selected 30 slices (=3.75cm) above the cranial reference point, and the caudal border was taken as the 176 slices (=22cm) below that. There was some variability in the cranial reference point, but since the axial FOV was sufficiently large, this did not lead to problems. Next, the central sagittal (reference) slice was estimated by the median slice for which the mean HUs exceeded 90% of the maximum (Fig. SF1.1c; blue arrow). The lateral FOV borders were then found by the slices ±96 slices from the selected central slice. Next, an anterior reference slice was found where the mean HU of center slice exceeded -950 (Fig. SF1.1d; blue arrow). This means that, in cases where a thermoplastic mask was used, the reference was more anterior than in cases where it was not used. From this reference, the anterior border was 1cm (10 voxels) posterior to the reference, and the posterior border was 176 slices from the anterior border. The FOV was then laterally divided in half (Fig. SF1.1e) to obtain the (unilateral) images used for training. CT HUs were then rescaled between [0-1]. Finally, images were checked such that all parotid gland voxels were contained well within the selected FOV and corrected where necessary.

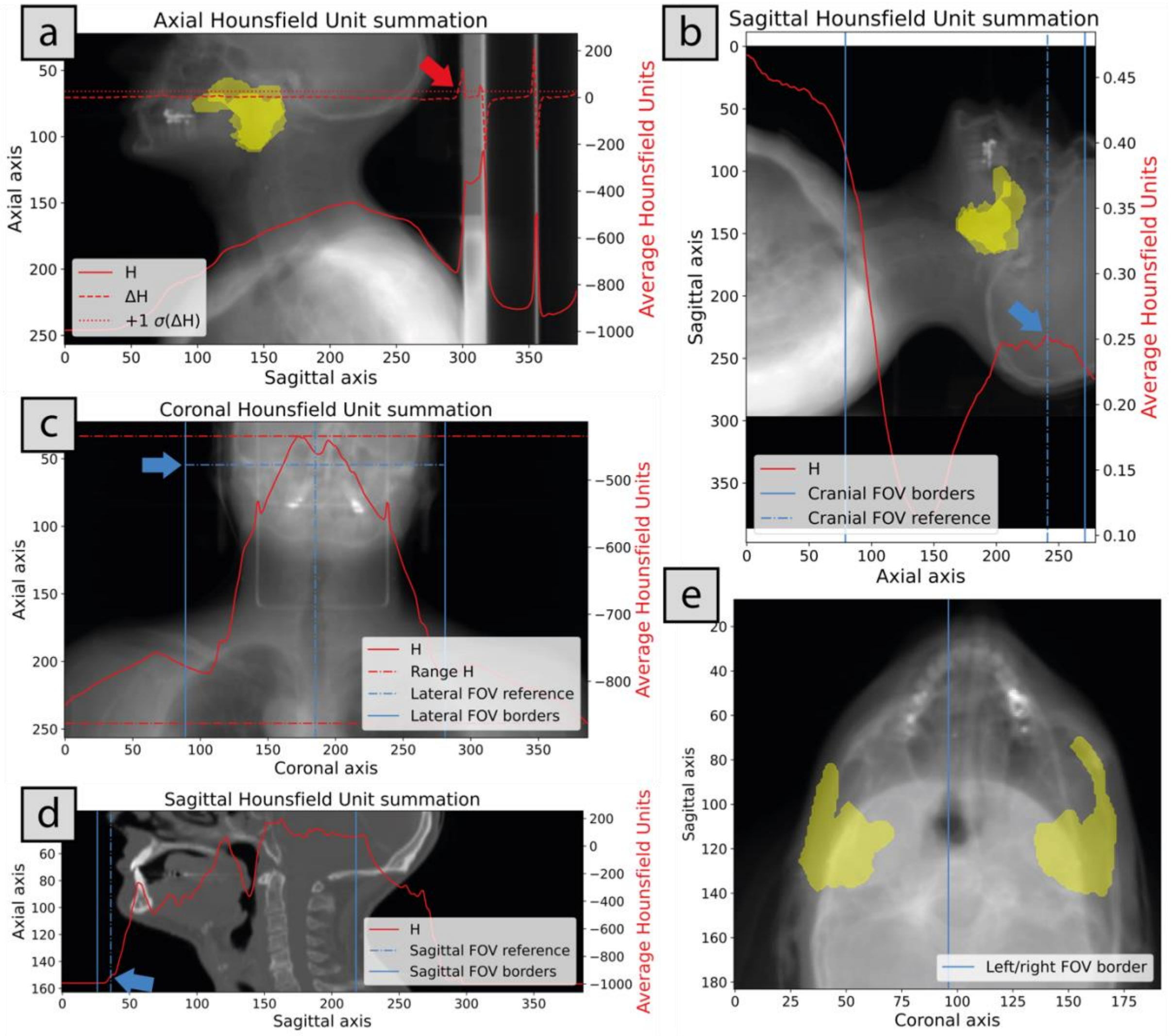


**Figure SF5:** Pre-processing overview. In sub-figures A-D, function H indicates the average Hounsfield Units (HUs) over the slice depicted on the x-axis. (a) Projection of CT HUs along the coronal axis. The couch was detected as the slice where the derivative of H exceeded one standard deviation (red arrow); (b) Projection CT in (a), but with the couch removed and rotated 90 degrees in the sagittal plane. The uppermost slice of the head was detected as the point in the face area where the HUs summed along the axial axis was maximal (blue arrow); (c) Projection of CT HUs along the sagittal axis. The central sagittal reference was estimated by the median slice for which H exceeded 90% of its maximum (blue arrow). This method was chosen because it was observed to make selection of the central slice more robust to rotations of the neck. Lateral FOV borders (blue solid lines) were then determined by the ±96 slices with respect to the reference central midpoint slice. (d) Anterior reference was selected where the mean HU of the center slice exceeded -950 (blue arrow); (E) Example of the selected 176x192x176 FOV. The yellow contours indicate projections of the parotid gland voxels in the clinical reference annotation.
