## Supplementary Material C for "Interpreting convolutional neural network explainability for head-and-neck cancer radiotherapy organ-at-risk segmentation"

### Supplementary Material C (Supplementary results)

#### Persistent homology PCA decomposition

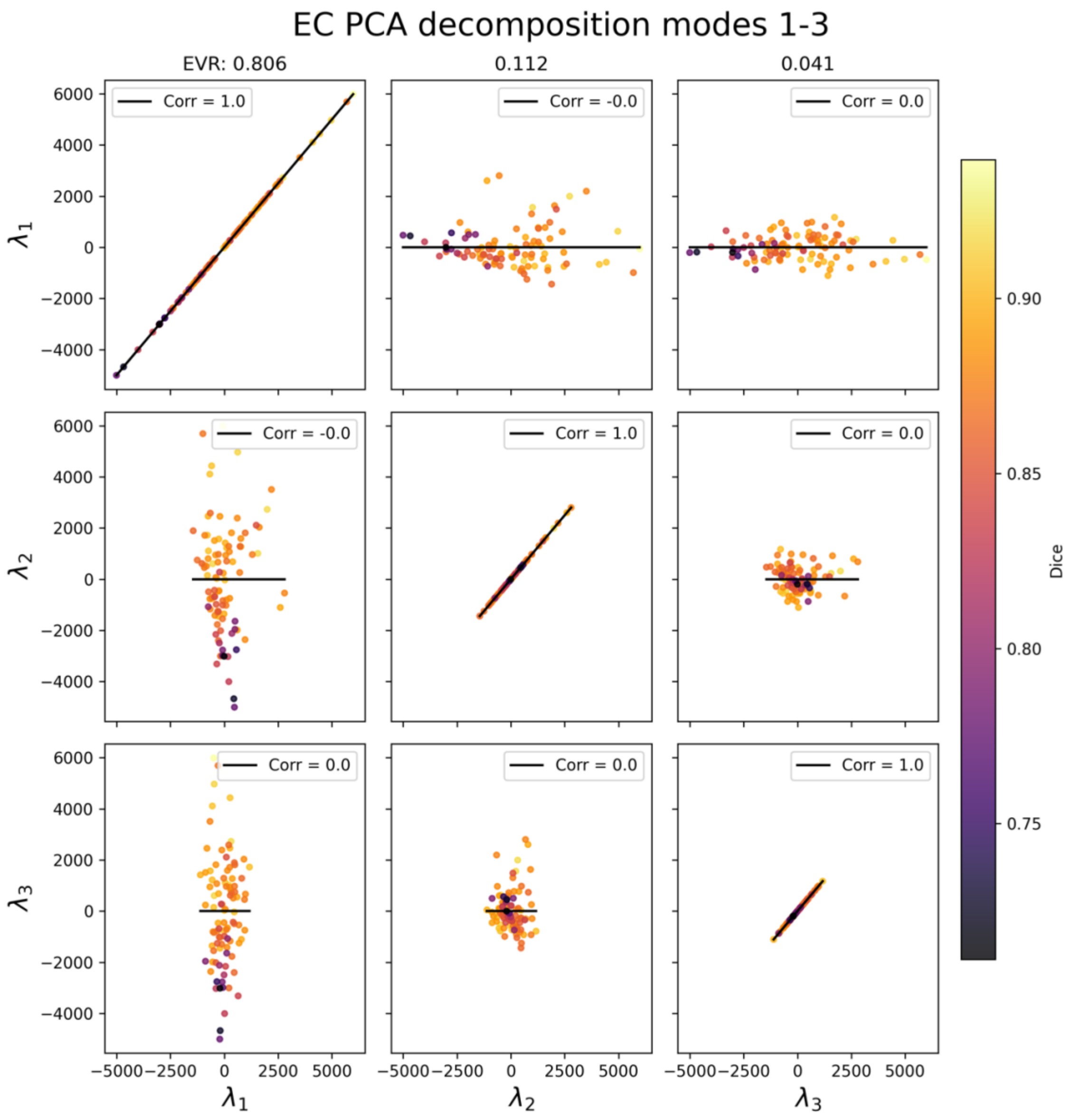


**Figure SF6:** PC modes 1-3 scatterplot matrix of GGCAM's Euler Characteristic (EC) curve. Abbreviations: PCA: Principal component analysis

### Attribution maps for supplementary cases

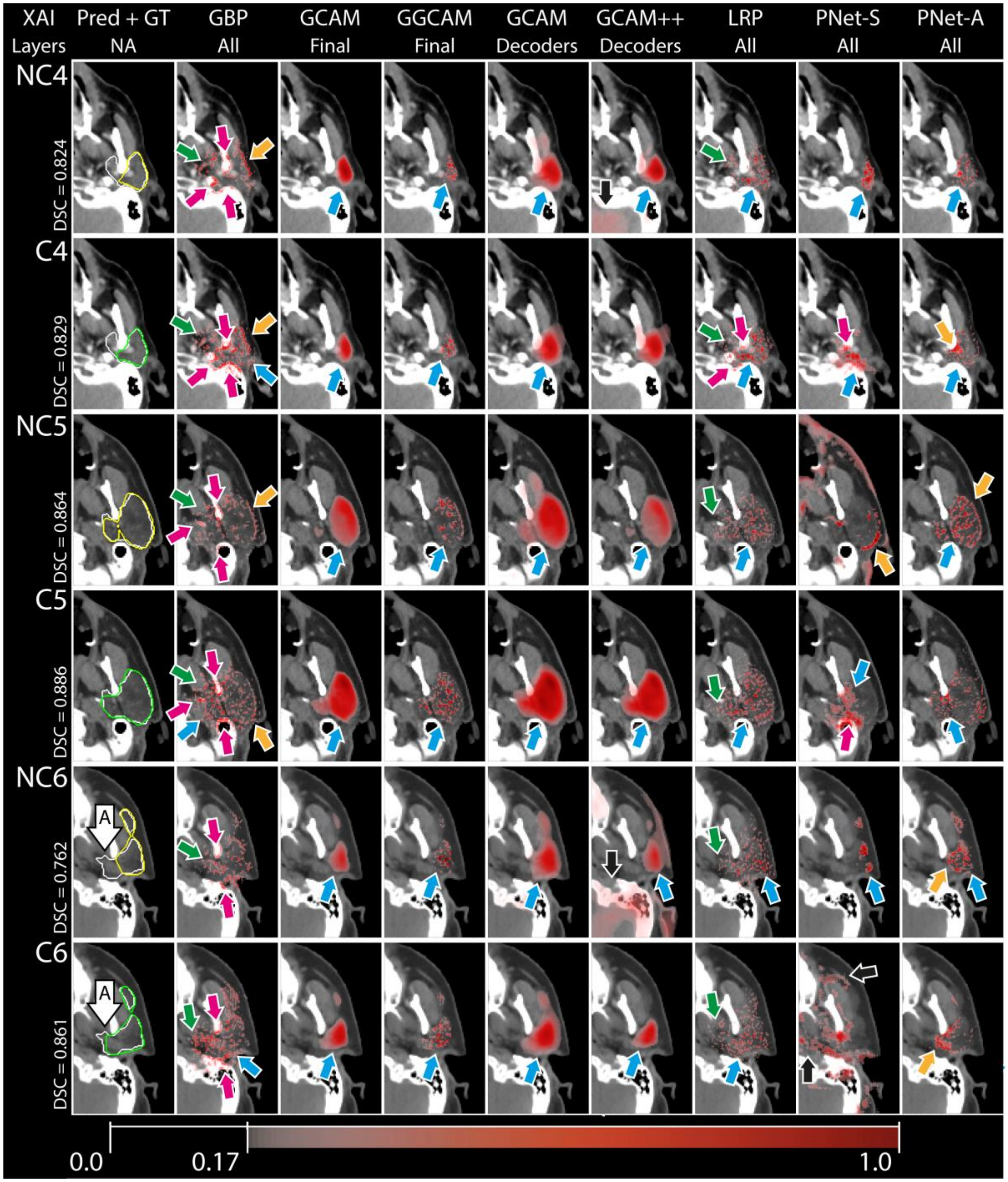


**Figure SF7** *Curated vs. non-curated model XAI comparisons cases 3-6.* Overview of XAI methods on clinical predictions comparing both non-curated (NC; yellow contour) and curated (C; green contour) CNNs. Arrows A indicate notable sites of improvement from curation in medial lobes. Three patients (C4-C6) were randomly sampled from each tercile of the curated DSC distribution. Abbreviations: GBP: guided backpropagation; GCAM: gradient class-activation mapping; GGCAM: guided GCAM; LRP: layer-wise relevance propagation; PNet: PatternNet; PNet-S: PNet signal; PNet-A: PNet attributions.

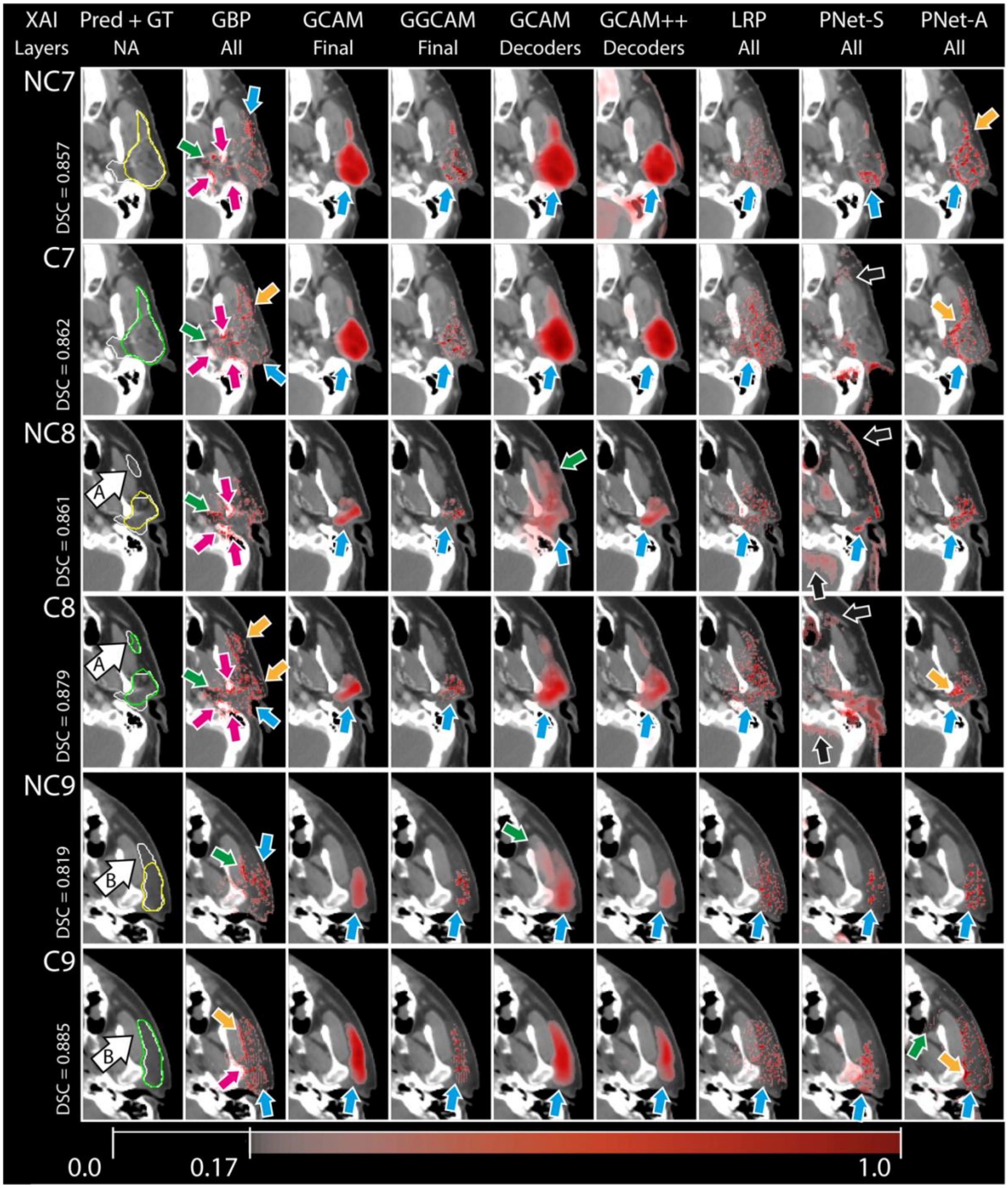

**Figure SF8** *Curated vs. non-curated model XAI comparisons cases 7-9.* Overview of XAI methods on clinical predictions comparing both non-curated (NC; yellow contour) and curated (C; green contour) CNNs. Arrows A and B indicate notable sites of improvement from curation in the accessory PG and anterior lobes, respectively. Three patients (7-9) were randomly sampled from each tercile of the curated DSC distribution. Abbreviations: GBP: guided backpropagation; GCAM: gradient class-activation mapping; GGCAM: guided GCAM; LRP: layer-wise relevance propagation; PNet: PatternNet; PNet-S: PNet signal; PNet-A: PNet attributions.

**Persistent homology for all XAI methods**

| | | GBP | GCAM | GGCAM | GCAM++ | LRP | PNet-S | PNet-A |
|---|---|---|---|---|---|---|---|---|
| **B0** | AUC | 0.65 | 0.58 | **0.94** | 0.42 | 0.58 | 0.61 | 0.84 |
| | Sp. $\rho$ | *0.06* | *-0.05* | *0.59* | *-0.15* | *0.03* | *0.11* | ***0.60*** |
| **B1** | AUC | 0.64 | 0.72 | **0.88** | 0.36 | 0.45 | 0.74 | 0.81 |
| | Sp. $\rho$ | *0.02* | *0.47* | *0.45* | *-0.12* | *0.06* | *0.31* | ***0.49*** |
| **B2** | AUC | 0.66 | 0.72 | 0.82 | 0.19 | 0.51 | 0.69 | **0.83** |
| | Sp. $\rho$ | *0.03* | *0.38* | *0.48* | *-0.19* | *0.11* | *0.23* | ***0.58*** |
| **EC** | AUC | 0.65 | 0.74 | **0.94** | 0.45 | 0.62 | 0.55 | 0.81 |
| | Sp. $\rho$ | *0.05* | *0.45* | ***0.54*** | *0.01* | *0.26* | *0.18* | ***0.54*** |

**Table ST2** Goodness of fit of the first PCA eigenvalue of topological features for DSC. B0-B2 refer to Betti-profiles 0-2 (i.e. numbers of connected components, circles, voids), whereas EC is the Euler characteristic profile (numbers of connected components – circles + voids). Best XAI method is denoted in bold. Abbreviations: AUC: Area-under-receiver-operating-characteristic curve; Sp. $\rho$: Spearman's rank correlation coefficient.

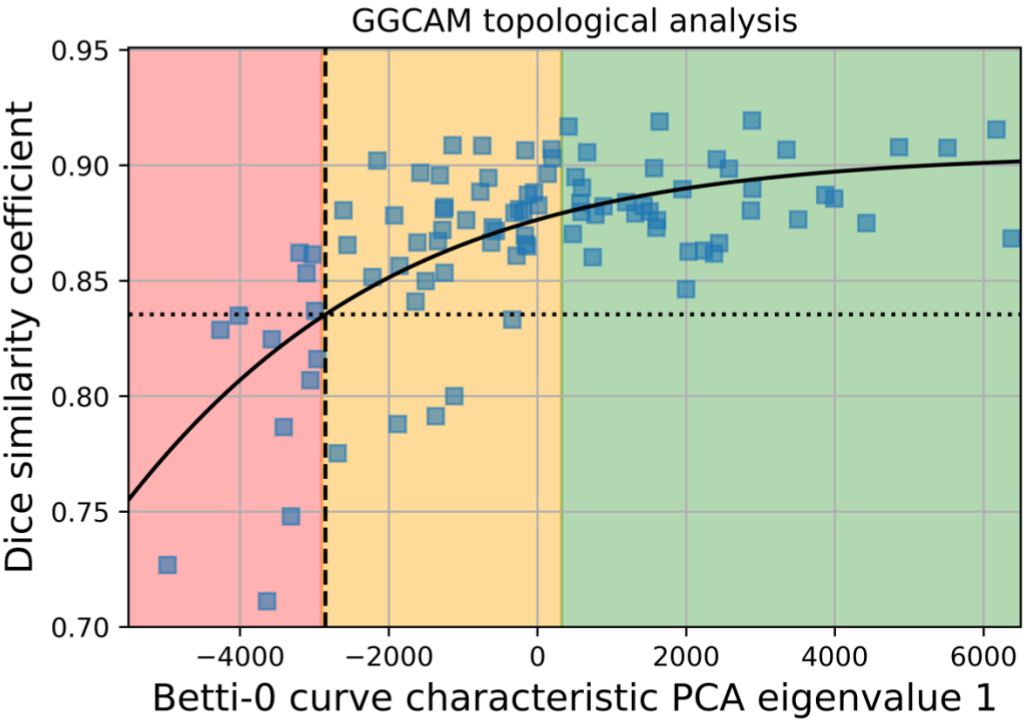


**Figure SF9** GGCAM's topological derivative for the Betti-0 profile.

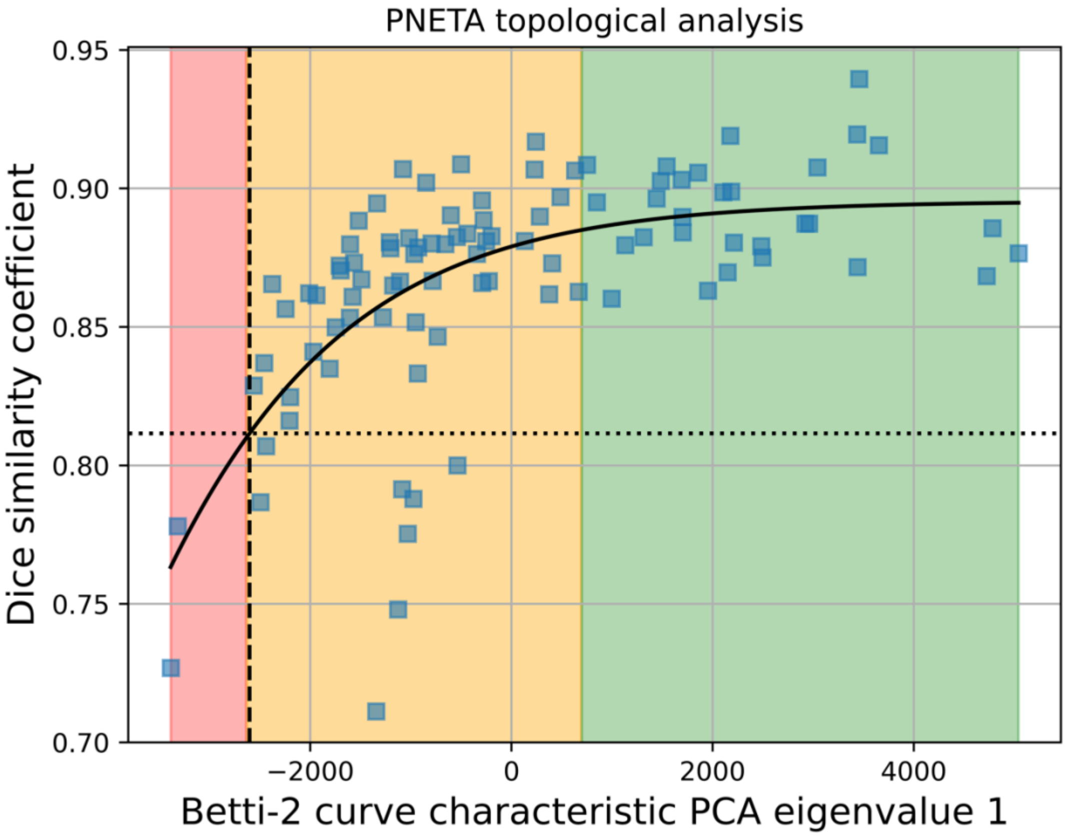


**Figure SF10** Topological derivative for the Betti-2 profile of PNet-A.
